# Validation of body surface colonic mapping against high resolution colonic manometry: a novel non-invasive tool for evaluation of colonic motility

**DOI:** 10.1101/2023.10.28.23297487

**Authors:** Sean HB Seo, Cameron I Wells, Tully Dickson, David Rowbotham, Armen Gharibans, Stefan Calder, Ian Bissett, Greg O’Grady, Jonathan C Erickson

## Abstract

Abnormal cyclic motor pattern (CMP) activity is implicated in colonic dysfunction, but the only tool to evaluate CMP activity, high-resolution colonic manometry (HRCM), remains expensive and not widely accessible. This study aimed to validate body surface colonic mapping (BSCM) through direct correlation with HRCM. Synchronous meal-test recordings were performed in asymptomatic participants with intact colons. A signal processing method for BSCM was developed to detect CMPs. Quantitative temporal analysis was performed comparing the meal responses and motility indices (MI). Spatial heat maps were also compared. Post-study questionnaire evaluated participants’ preference and comfort/distress experienced from either test. 11 participants were recruited and 7 had successful synchronous recordings (5 females/2 males; median age: 50 years [range: 38-63]). The best-correlating MI temporal analyses achieved a high degree of agreement (median Pearson correlation coefficient (*R*_*p*_) value: 0.69; range: 0.47 - 0.77). HRCM and BSCM meal response start and end times (*R*_*p*_ = 0.998 and 0.83; both *p <* 0.05) and durations (*R*_*p*_ = 0.85; *p* = 0.03) were similar. Heat maps demonstrated good spatial agreement. BSCM is the first non-invasive method to be validated by demonstrating a direct spatio-temporal correlation to manometry in evaluating colonic motility.

## Introduction

Disorders of lower gastrointestinal (GI) function affect 10-33% of the global population, and significantly impact quality of life^1–4^. Costs of investigations and treatments impose a substantial burden to healthcare systems^5–7^. In the past decade, translational studies using high-resolution colonic manometry (HRCM) have found that alterations in colonic cyclic motor pattern (CMP) activity are implicated in a diverse range of functional bowel disorders^8–13^. These studies have had a pivotal role in elucidating our understanding of colonic motility and function, and HRCM has become the ‘gold standard’ tool for researching colonic motility^14,15^. Modern HR manometers have sensors at up to 1 cm resolution, allowing detection of shorter propagating sequences (i.e., CMPs) than previous low-resolution devices^16^. When paired with X-ray imaging, propagating events can be localized to an area along the colon to enable spatio-temporal correlation^10,17–19^.

However, while HRCM is a valuable research tool, it has not been widely adopted for clinical use due to its limited availability, invasiveness, cost, and the complexity of analysis. Conventional diagnostic tests for colonic functional disorders that are widely available are principally transit studies and do not directly assess motility patterns, limiting the depth of pathophysiological data that can be assessed^20^. The lack of an accessible tool to identify physiological biomarkers has led to the wide use of symptom-based definitions for functional disorders^21–23^ which impedes therapeutic advances. Understanding the motility phenotypes of the colon that correlate with specific dysfunctions will be essential to develop targeted and individualized therapies in the future.

Body surface electrical mapping technologies have recently emerged as a novel approach to evaluating GI electrophysiology and function^24,25^. These techniques are non-invasive, easy to implement, and have recently achieved regulatory approvals in gastric conditions^26,27^, paving the way for a similar technical platform for non-invasively evaluating clinically relevant biomarkers in colonic (dys-)function. For the stomach, non-invasive body surface gastric mapping (BSGM) has been validated against invasive high resolution (HR) serosal mapping^24,26,28^. For the colon, animal and human studies using internal and/or external electrical sensors have detected rhythmic colonic electrical activities of congruent frequency; however there have been no studies directly correlating outputs from non-invasive methods to manometry^29,30^. Evaluating colonic activity can also be complex relative to the stomach, due to larger anatomical variations, intermittent activity profiles, a more diverse frequency range, and potential for multiple synchronous active regions with independent CMP characteristics^31^.

In order to advance the clinical translation of non-invasive colonic mapping, the current study aimed to validate the performance of Body Surface Colonic Mapping (BSCM), an automated signal processing method for extracting the colonic signal from body surface electrical recordings, against simultaneous HRCM recordings in healthy individuals. Validated BSCM methodology could enable translational research in colonic function to be much more acceptable to participants, less costly for researchers, and may accelerate the identification of clinically significant biomarkers of colonic function. The results show that BSCM can detect CMP activity in the colon, quantify meal responses, and spatially locate hotspots of CMP activity with a high level of correlation to HRCM.

## Methods

Ethical approval was obtained from Auckland Health Research Ethics Committee (AHREC); AH1287. This study was conducted in accordance with the ethical principles outlined in the Helsinki Declaration. All participants provided informed consent which included the use of non-identifying photographs in publication of the study.

### Eligibility criteria

Adult patients (aged 18 years or above) were recruited who were planned for elective colonoscopy for surveillance or for indications correlated with a low likelihood of underlying diverticular disease or colorectal cancer in Auckland City Hospital, Auckland, New Zealand. Patients were excluded if they were pregnant, had a history of previous colonic resection, diagnosed GI motility disorders or comorbidities that affect bowel motility/function (Parkinson’s disease, thyroid function diseases, and diabetes mellitus), or if they had a regular prescription for agents affecting bowel function (including, but not exclusive to, loperamide, prokinetics, metformin, laxatives, and opioids).

### Study protocol

High-resolution colonic manometry A fiber-optic manometry catheter with 72 sensors at 1 cm intervals was used^32^. All participants agreed to have placement in the same procedure as the elective colonoscopy. Following the clinical colonoscopy, a re-entry was made with an endoscopic grasper holding a nylon loop which had been placed at the proximal tip of the manometer. The catheter was placed in the maximum possible extent in the right colon. At extent, the nylon loop was endo-clipped to the colonic mucosa to keep the catheter in place and the colonoscope was gently removed. The manometer was fixed in position with an adhesive dressing to the right buttock to prevent unintended withdrawal during the recording period. A spectral interrogator acquisition unit was connected to the catheter to record the data (FBG-scan 804; FOS&S, Geel, Belgium). (See Fig. 1).

**Figure 1.**
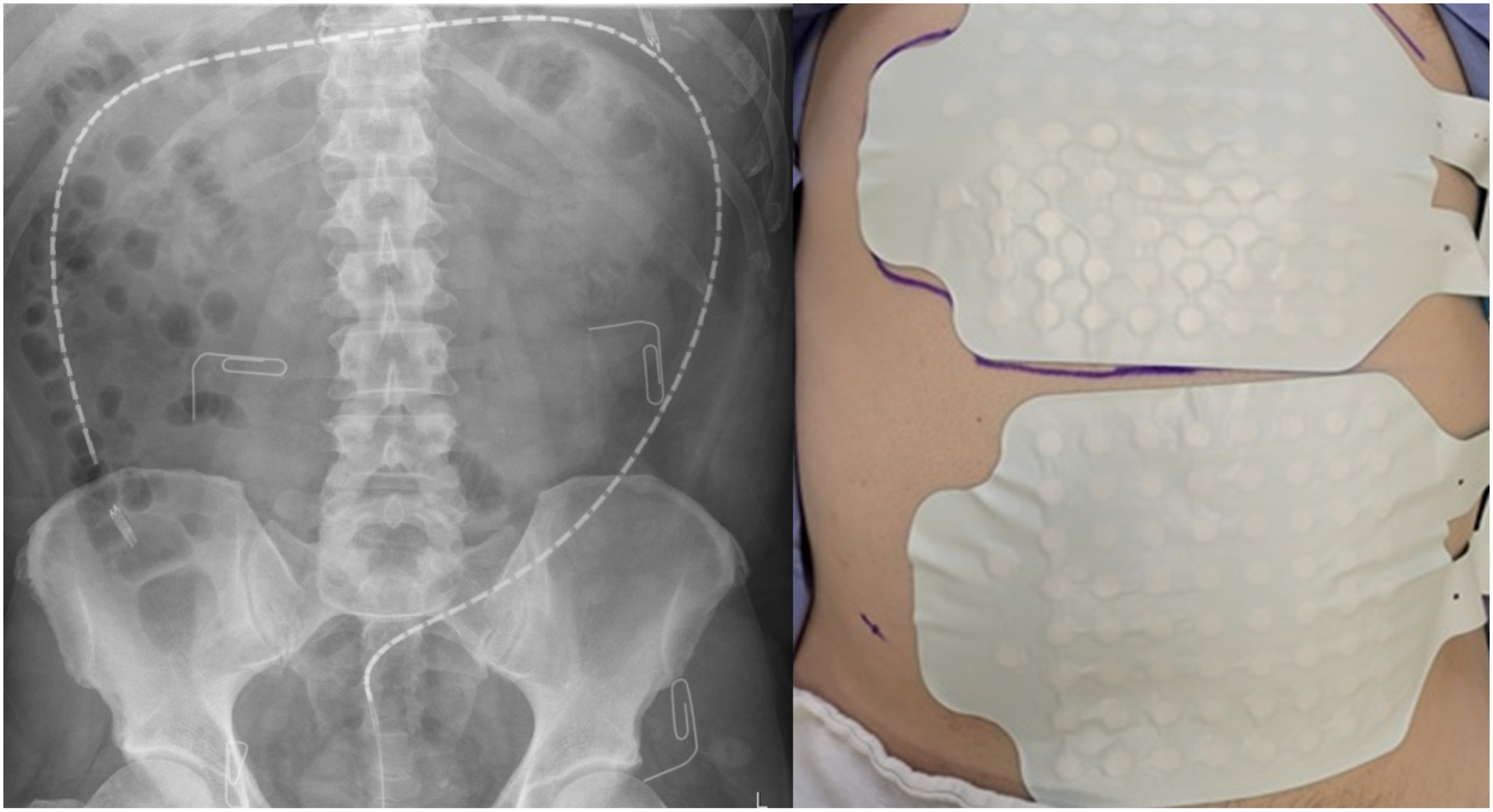
High resolution fiber-optic colonic manometer and body surface mapping (BSM) array placement. **LEFT**: 72cm manometer with a sensor for each 1 cm was clipped with multiple endoclips to the mucosa of the colon. The paperclips demarcate the area covered by the lower 8 × 8 electrode grid of the array. **RIGHT**: BSCM array is affixed to abdominal skin. Ground and reference electrodes are located on the small extension flap of the arrays on the right (anatomical) side of the participant.

### Body surface colonic mapping

The BSCM electrical recordings were acquired using the Alimetry body surface mapping hardware (Alimetry Ltd, Auckland, New Zealand) with an 8 *×* 8 stretchable electronics adhesive array with pre-gelled Ag/AgCl electrodes at 2 cm spacing; area 225 cm^2^ (see Fig. 1)^26^. Prior to array placement, abdominal hair was clipped, and the skin was prepared with an exfoliant (NuPrep; Weaver, Aurora, CO) to facilitate low impedance electrical contact. The array was positioned on the lower abdomen primarily to capture electrical activity arising from the rectosigmoid junction and the left colon^33,34^.

### Experimental protocol

On the morning of the experiment, informed consent was obtained. All participants completed the standard regimen of the bowel purgative agent Glycoprep-C (Fresenius Kabi, Australia). The choice and dose of procedural medications (anesthetic and analgesia) were decided by the endoscopist/anesthetist. Smooth muscle relaxants, such as hyoscine, were not administered. Following placement, the manometer was connected to the acquisition unit, and the BSCM array was placed on the abdomen and connected to the Alimetry Reader. Participants underwent a plain abdominal x-ray with the corners of the array marked with radio-opaque clips to spatially register the position of the manometer in relation to the body surface array (see Fig. 1). Concurrent data acquisition was performed for three to four hours (one hour pre-meal period, two to three hours post-meal). During the recording, participants were asked to lie in a reclining chair/bed positioned 20-30 degrees from horizontal. The standardized meal included a 232-kcal nutrient drink (230 mL Ensure; Abbott Nutrition, IL, USA) and an oatmeal energy bar (250 kcal with 5 g fat, 45 g carbohydrate, 10 g protein, 7 g fiber; Clif Bar & Company, CA, USA). Participants were given 10 minutes to complete the meal. At the end of the study participants were asked to fill out an electronic questionnaire pertaining to the comparative experience of HRCM and BSCM. Likert scales from 1 to 10 were used for perceptions of discomfort/pain and usability.

### Data analysis

The protocol included a 10 minute settling period, which was not analyzed. The earlier end of data acquisition of either the HRCM or BSCM device was deemed the end of time of the experiment.

### HRCM analysis

Primary analysis was performed using PlotHRM (Flinders University, Adelaide, Australia). Markers were placed manually on the consecutive peaks of propagating or simultaneous events. Criteria for motor events were pressurizations of 5 mmHg or greater across four or more consecutive channels (i.e., 4 cm or longer)^8,17,33^. All pressure events in multiple (2 or greater) within 1 minute of each other were marked and included in the analysis. Custom MATLAB R2022b (MathWorks, Natick, Massachusetts, USA) software was used for further analysis. Amplitude (in units of mmHg) was defined, following previous studies, as the average of the peak pressures noted in every channel involved in a propagating pressure wave^10,35^. Length of a CMP (units of cm) was defined as the distance between the first and last manometer sensors along the same marked sequence—e.g., 5 consecutive sensors involved making a CMP 4 cm in length. The instantaneous rate of CMPs was determined from the number of propagating events occurring within a sliding 2 minute window. To prevent double counting, a single propagating event was time-stamped at its mid-point. HRCM frequency was analyzed using two separate methods. Intrinsic frequency was defined as the rate of CMP activity detected on each individual manometer sensor, independent of other sensor’s data, thus representing the electrophysiological rate of region-specific CMP activity. Sequential frequency was defined using the time interval of 2 successive events marked array-wide on any sensor, thus representing the rate of multifocal CMP activities along the entire colon superposing at the body surface. When only one region is active, the sequential frequency equals the intrinsic frequency. In this paper, HRCM frequency refers to the intrinsic frequency unless stated otherwise.

### BSCM analysis: preprocessing methods

An optimized BSCM signal processing method was developed on the foundations of a previous proof of concept study that used continuous wavelet transformation (CWT) analysis to characterize the electrical activities occurring within a set frequency bandwidth^29^. Guided by HRCM frequency analysis and other literature, it became prudent to further develop the signal processing pipeline to be capable of monitoring a wide and dynamic frequency range at the body surface. Initially, three frequency bandwidth filters were used to cover the colonic frequency ranges previously stated in literature^31,36^ and observed in the HRCM results in this study; low (0.6-6 cpm), high (5-12 cpm) and wide (0.12-12 cpm). Subsequently, frequency ranges were fine-tuned on an empirical basis. For each of 10 total frequency bands tested, we explored whether application of common mode re-referencing (CMR) and linear minimum mean square estimation (LMMSE) artifact reduction improved the correlation with HRCM MI. In total, we explored a parameter space consisting of 40 total preprocessing combinations.

Raw BSCM recording were processed as follows:

1. Remove baseline wander using a moving median filter with a 30 s window.
2. If selected, reduce motion artifacts using LMMSE filter with 30 s averaging window and adaptive noise threshold window of 300 s^37,38^.
3. Attenuate noise and interference while isolating various putative colonic components using a Butterworth filter (2nd order; zero phase). We tested passbands (in units of cpm): 0.12-12, 0.6-6, 0.6-12, 2-8, 3-10, 4-10, 4-12, 5-8, 5-12, and 8-12 (see also Fig 3b).
4. If selected, apply common mode re-referencing (CMR) following the PREP pipeline^39^. CMR was applied using only channels with good signal quality, based on criteria of low electrical impedance (≤ 500 kΩ); low noise but not ‘dead channel’ amplitude (1 - 4000 *μV*); sufficient spectral correlation to other channels (correlation ≥ 0.3). Only ‘good’ channels meeting all of the above criteria were included for the remaining analyses; ‘bad’ channels were excluded.
5. An additional stage of large transient reduction was applied based on soft-thresholding. The soft-thresholding coefficient was determined as:

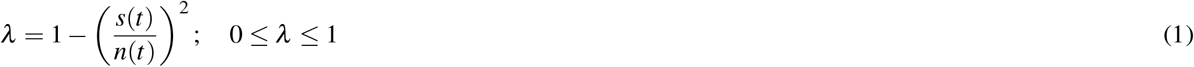

Here, *s*(*t*) is the magnitude of the Hilbert Transform of the signal resulting from steps 1-4 above, smoothed over a 300 s window. It’s ratio relative to *n*(*t*) = *median*(*s*(*t*)) + *k ×MAD*[*s*(*t*)] sets the threshold level, where *MAD* is the median of the absolute deviation, an estimate of the signal variance. We set *k* = 5, a choice known empirically to work well, and a maximum value for *n*(*t*) = 500*μ*V. Note that colonic signal amplitudes are expected to be in the range of 50 − 200*μV*, such that non-artifact corrupted segments of the signal amplitude are only modestly attenuated by ≈ 1 − 16%, while larger transients are strongly reduced.

### Motility indices

#### HRCM MI

In this study, we focused on aspects of CMPs that should be detectable from the body surface including temporal activation and localization of the regions of CMP activity. Specific characteristics of any individual motility events, such as direction and velocity of a CMP, were judged less likely to be relatable to BSCM electrophysiological signals owing to complex orientation and geometry of the colon and the spatial volume conductor effect. Thus, we defined the HRCM motility index (*MI*) representing the summative CMP activity as a product of three metrics: number of CMPs per unit time (*N*_*CMP*_); mean amplitude (*A*) and distance of propagation (*L*):

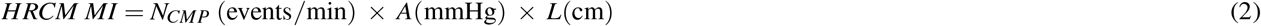

#### BSCM MI

BSCM MI was derived from Continuous Wavelet Transform (CWT) analysis in a 3-step process. First, for each channel the CWT spectrogram was computed as previously described^29^. Note the magnitude of CWT coefficients represent the amplitude of the signal at each frequency (scale) and time point. Second, the array-wide spectrogram was computed by averaging CWT spectrograms from all individual-channels. Finally, BSCM MI was computed as the mean of the top 10% wavelet coefficient values at each time point. We termed this the ‘CWTmaxval’ method. Because the preprocessing stage accurately rejects artifacts and noise components, CWTmaxval is a robust metric quantifying the colonic motility over time, which may exhibit a variable dominant frequency.

### Comparative analysis/outcomes: temporal analysis

#### Meal response

Meal response start, end and duration times were assessed independently by 2 researchers (SHBS and JE) and any disagreements were resolved by a third reviewer (CW). Meal responses start time was the earliest time at which MI rose above the baseline level for longer than 10 minutes. The point at which the MI returned to baseline levels, sustained for the ensuing 10 minutes or long, was the end time of the meal response.

#### Motility index correlation

Correlation analyses evaluated which BSCM MI result for each of 40 preprocessing parameter sets (10 filter bandwidths *×* 2 artifact reduction settings (on/off) *×* 2 CMR settings (on/off)) best represented the CMP activity as apprised by manometry. HRCM MI and BSCM MI were smoothed over a 5 minute window, chosen to reduce noise while preserving temporal dynamics characteristic of episodic CMPs observed in this study cohort.

### Spatial correlation

The abdominal x-ray images were used to register the approximate location and orientation of the manometer and the body surface array. To spatially compare the dominant region(s) of CMP activity, the experiment time course was divided into manually identified epochs that we considered to be major phases of the meal response; pre-meal, meal response, post meal response (quiescence) and secondary active segments (if any) as identified from HRCM analysis. Spatiotemporal activity in each of these time periods were compared using HRCM activity maps to BSCM heatmaps, visualizing the most active zones detected in each, respectively. Qualitative analyses assessed the synchronicity and general location of dynamic spatial ‘hot spots’ during the meal response epochs.

#### HRCM

HRCM spatiotemporal activity maps were generated in each temporal epoch using a two-step process as follows:

1. The cumulative mean activity for each sensor was computed as 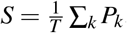, where *P*_*k*_ denotes pressure amplitude of the *k*th marked event within the time window of duration *T*.
2. Bivariate kernel smoothing was applied with a bandwidth equal to ≈ 1.5% of the mean scale of the image (in pixels). This last step was done primarily to match the approximate diameter of the colon in the X-ray image.

#### BSCM

For each time epoch, the BSCM heatmap was rendered follows:

1. For each good channel, the BSCM MI was scaled to have unit area under the curve—i.e., *BSCM MI*(*t*)*/* ∑_*t*_ *BSCM MI*(*t*). Normalization helps compensate for variable source to sensor distance across the array, emphasizing overall changes in activity^29^.
2. The mean BSCM MI within a defined temporal epoch was computed for each electrode.
3. To fill gaps in the map where bad electrodes existed, inverse cubic weighting interpolation (distance scale = 2 pixels) temporal weighting was applied with a radius of 2*×* electrode distance.
4. To reduce the effect of any remaining outliers in the heat map, a thin plate spline was applied (smoothing parameter = 0.5).
5. The 8 *×* 8 grid of values was up-sampled by a factor of 10 for a clearer visualization.
6. Global color scale limits were set using the 25-99 percentile of values in the heat map. These values were empirically determined to be a good compromise between sufficiently rendering detail in a single map versus highlighting large contrasts in activity that may exist across epochs (e.g., quiescent vs active).

### Statistical analysis

Student’s *t*-test was performed to compare the difference between the meal response outcomes measured from the manometry data and BSCM data. The null hypothesis was that there was no difference between the two datasets. Pearson’s correlation coefficients were used to assess the strength of correlations between the motility index (*MI*) outcomes of the recordings. A p-value of *<* 0.05 was deemed to show statistical significance between two datasets. Pearson coefficient *>* 0.5 was considered to be ‘good’ or substantial agreement between MI traces. GraphPad Prism version 9.5.1 (GraphPad Software, San Diego, California USA) and MATLAB R2022b (MathWorks, Natick, Massachusetts, USA) were used for statistical analyses and production of figures.

## Results

### Participants information

A total of 11 participants were recruited with a median age of 50 (range: 30 to 69) and majority were female (9:2). Simultaneous recordings for analysis were successfully achieved in 7 participants whose median BMI was 25.6 (range: 22.3 to 31.3). Recordings from 4 subjects were excluded from further analysis due to: insufficient overlapping data due to a delayed x-ray and loss of over half of the manometry data due to setting/connection error (1), manometer insertion failure (1), manometer technical failure and inadequate BSCM data (1), and participant early withdrawal (1). One participant had a pre-arranged propofol anesthetic and one participant declined both sedation and analgesia. Indications for colonoscopies were family history (3), anemia (2) and mild GI symptoms at the time of referral which had settled completely by the time of investigation (2). All other participants received a combination of intravenous midazolam and fentanyl, and none were administered hyoscine or other antispasmodic medications (see Table 1).

**Table 1.** Demographics of participants.

| Subj | Age | Sex | Ethnicity | BMI | Clinical Indication | Sedation |
| --- | --- | --- | --- | --- | --- | --- |
| 1 | 35–39 | F | Middle Eastern | 22.7 | Iron deficiency anemia and rectal outlet bleeding | Propofol |
| 2 | 50–54 | M | NZ European | 25.6 | Intermittent abdominal discomfort and dyspepsia | Midazolam + Fentanyl |
| 3 | 60–64 | F | NZ European | 24.4 | Family history of bowel cancer and previous polyps | Midazolam + Fentanyl |
| 4 | 60–64 | F | NZ European | 22.3 | Altered bowel habit and family history of bowel cancer | Midazolam + Fentanyl |
| 5 | 40–44 | F | Other European | 25.8 | Bloating/epigastric pain | Midazolam + Fentanyl |
| 6 | 50–54 | F | NZ European | 31.3 | Anemia | Midazolam + Fentanyl |
| 7 | 50–54 | M | NZ European | 31.0 | Lynch syndrome - regular surveillance | Nil |

The median recording duration was 211 mins (range: 50-239). The first two of the analyzed cases had irregular pre-meal periods of 23 and 102 minutes due to being called for the X-ray imaging. In the subsequent studies, X-ray imaging was performed pre-recording, facilitating uninterrupted synchronous recordings of pre-meal (60 mins) and post-meal periods.

### Data summary

Synchronously recorded BSCM and HRCM data from the 7 subjects with successful recordings were analyzed. A meal response occurred in all, with timings and duration observed to be highly concordant between HRCM and BSCM measurement modes. Temporal analysis of BSCM and HRCM motility indices demonstrated a high level of correlation (median Pearson *r* = 0.69; 0.47 − 0.77) using the subject-specific optimal signal processing method for each case. Collectively across the cohort, the frequency bandwidth range 4-10 cycles per minute (CMR = off; LMMSE artifact reduction = on) was identified as the best single signal processing method (out of 40 possible) to achieve the highest correlation to the manometry data (median Pearson *r* = 0.63; 0.43 − 0.69). Spatial analyses showed good agreement of the regions of CMP activation during the active and quiescent epochs. Participants’ experience was significantly more positive with BSCM compared with HRCM; all participants unanimously preferred BSCM to HRCM and less discomfort was reported with BSCM (HRCM: median 7.5/10; range 2-9 vs. BSCM: median of 1/10; range 1-5; *p* = 0.0005).

### HRCM frequency and best correlating BSCM frequency bandwidths

Colonic intrinsic frequency was dynamic and generally rose to higher frequencies following the meal. For example, during subject 4’s meal response CMPs were mainly above 6 cpm and showed 2-3 minute bursts of high frequency activity in the rectosigmoid region up to 12 cpm. Subjects 3, 6 and 7 also had dynamic rises in their CMP frequency during the post meal period. Only subjects 1, 2 and 5 continued to exhibit low frequency (≈ 2 − 4 cpm) CMPs in their post meal periods (see Fig. 2). These findings are all within the ranges of normal CMP physiology stated in the literature^31,33^. Low frequency CMPs (2-4 cpm) were the dominant intrinsic frequency type (2-4 cpm activity: 46.4%, 4-10 cpm activity: 33.3%; *p* = 0.036). However, the sequential frequency range analysis showed the opposite result; a significantly higher proportion of CMP activity incurred within the higher frequency bandwidth (4-10 cpm activity: 47.0% vs 2-4 cpm activity: 26.5%; *p* = 0.0069) (see Supplementary information: Table S1). The cohort-wide optimally correlating preprocessing settings were determined to be: frequency range = 4-10 cpm, LMMSE = on; CMR = off (see Motility index match below for details).

**Figure 2.**
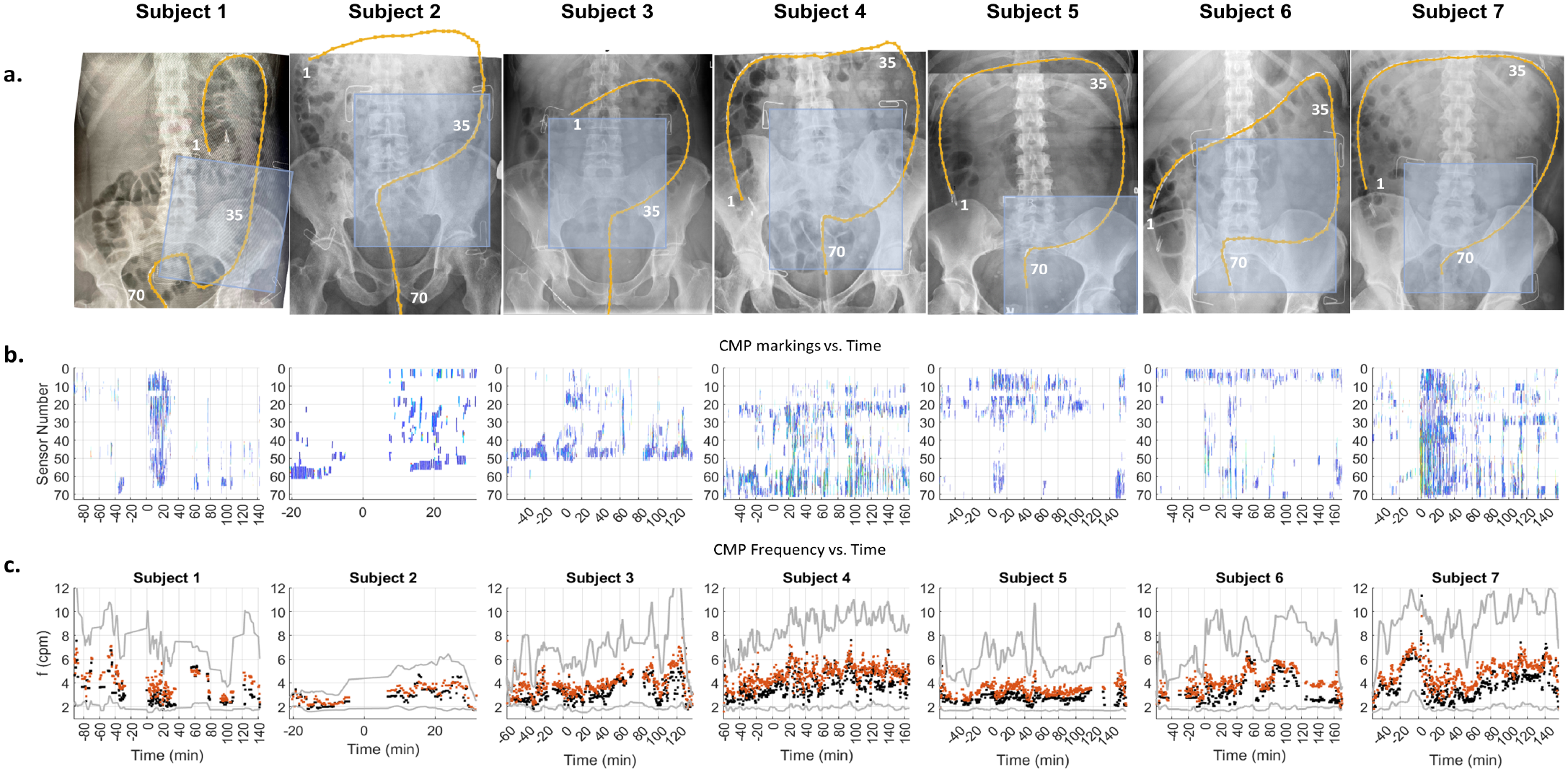
HRCM placement, manual markings, and dynamic frequency of CMPs: **(a)** X-ray images of each participant showing the extent the manometer (orange line) reached. Sensor numbers 1, 35, and 70 are annotated. **(b)** Manually marked CMP events graphically stacked with the most proximal sensor located at the top of the vertical axis (sensor number 1). The time axis is displayed with the meal start aligned at t = 0. CMP events are color coded according to intrinsic frequency (linear scale of 1.5 cpm [deep blue] to 10.5 cpm [red]) to highlight the dynamic frequency changes. Densely marked (pacemaker) regions of CMP activity can also be appreciated and anatomically localized which can then be used to develop a spatial ‘heatmap’ along the length of the manometer. Multi-focal activity can be observed as clusters of CMP marks that are vertically discrete with minimal activity occurring in the sensors separating them. **(c)** Using the data from B, frequency time course maps were developed. Orange dots indicate the mean frequency of the epoch, black is the median frequency and the gray lines indicate the 10-90 percentile range.

### Motility index correlation

Overall, the CMP activity for every subject was successfully identified and correlated well between manometry and body surface measurement modes (see Fig. 3). Even highly dynamic motility was observed to be highly correlated. For example, subjects 5, 8, 9 and 11 all had secondary activities (rise in CMP activity after the primary meal response had completed per this study’s definition of meal response) that were closely concordant in BSCM and HRCM MI traces versus time.

**Figure 3.**
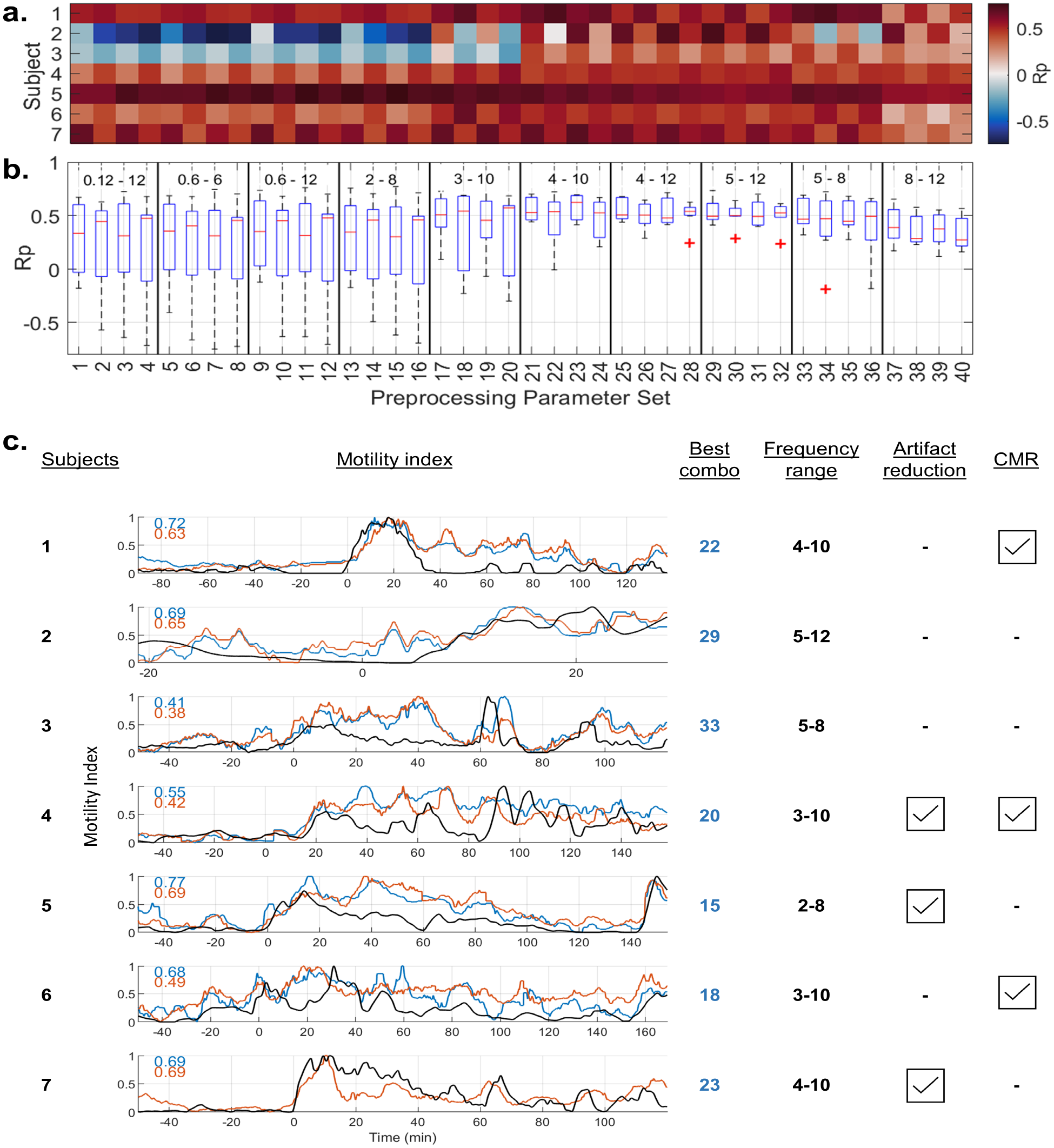
Motility index temporal correlation grid and time course of the best matching signal processing pipelines: **(a)** BSCM-HRCM MI correlation values across 40 different combinations (4 columns per frequency bandwidth tested) of signal processing performed for each of 7 subjects. The sets of 4 preprocessing filter combinations within a specified frequency band are specified as (artifact reduction, CMR) = (off, off); (off, on); (on, off); (on, on). Pseudocolor indicates values of Pearson correlation coefficient (*R*_*p*_). **(b)** Box plot statistical summary computed across all subjects for each of 40 individual preprocessing parameter sets. Combination 23 (frequency 4 - 10 cpm, artifact reduction on, CMR off) yielded the cohort-wide best overall performance. **(c)** Motility index vs. time: Black trace = HRCM MI, blue trace = maximum correlating BSCM MI; and red-orange trace = cohort-wide best overall preprocessing parameter set (number 23). Meal times are aligned at t = 0 min. Every subject had a different preprocessing parameter set achieving maximal correlation, but there was a strong trend toward higher frequency range filtering.

The optimal (maximal correlation) signal processing parameter set varied across subjects (see Fig. 3). Artifact reduction was favorable in 3 out of 7 cases, similarly CMR results were just as divided. While searching for the best overall signal processing combination, out of the possible 40, the higher frequency filter bands, with the lower cutoff *geq*4 cpm, were significantly better in 4 out of 7 cases. Frequency bands with lower cutoffs ≤ 3 cpm resulted in poor performance (negative correlation values) for 2 subjects (2 and 3), and ≤ 2 cpm cutoff additionally yielded reduced correlation for 2 more subjects (4 and 6). Across the 7 recordings, the signal processing combination with frequency bandwidth 4 to 10 cpm; artifact reduction on, CMR off (combination number 23 in Fig. 3) offered the best cohort-wide performance (Pearson *r* mean *±* std = 0.56 *±* 0.13; median = 0.63; range = 0.38 - 0.69) (see Fig. 3b). The motility index correlation using individually optimal signal processing pipelines (see Fig. 3a) led to modestly higher levels of correlation (Pearson *r* mean *±* std = 0.64 *±* 0.12; median = 0.69; range = 0.41 - 0.77).

### Meal response match

Meal responses were successfully identified in all 7 subjects. All measured endpoints correlated strongly between manometry and body surface data (Figures 4a and 4b). The meal response start time had a very strong correlation (Pearson *r* = 0.998; *p <* 0.0001). The meal response end time (Pearson *r* = 0.83, *p* = 0.041) and meal response duration (Pearson *r* = 0.85; *p* = 0.03) between HRCM and BSCM were also strongly correlated. Subject 2 was not included in the end time and duration analyses as the experiment ended early in the initial period of the meal response (53 minutes total recording duration). As illustrated in Figure 3, meal response activities started at the beginning of the meal ingestion period, aside from subject 4, whose primary meal related activity rose just after the meal was completed. The correlation for meal response end time and duration correlation was weakened by subject 1, an outlier. At the end of subject 1’s meal response on manometry, BSCM also shows a drop in the MI (approximately halved), however, the MI level is maintained above the baseline for another hour (see Figure 3c). The BSCM spatial analysis of subject 1 suggests that the meal response continues in a region not reached by manometry—in the proximal/right colon—which may account for the discrepancy between HRCM and BSCM (see Supplementary Information: Figure S1).

**Figure 4.**
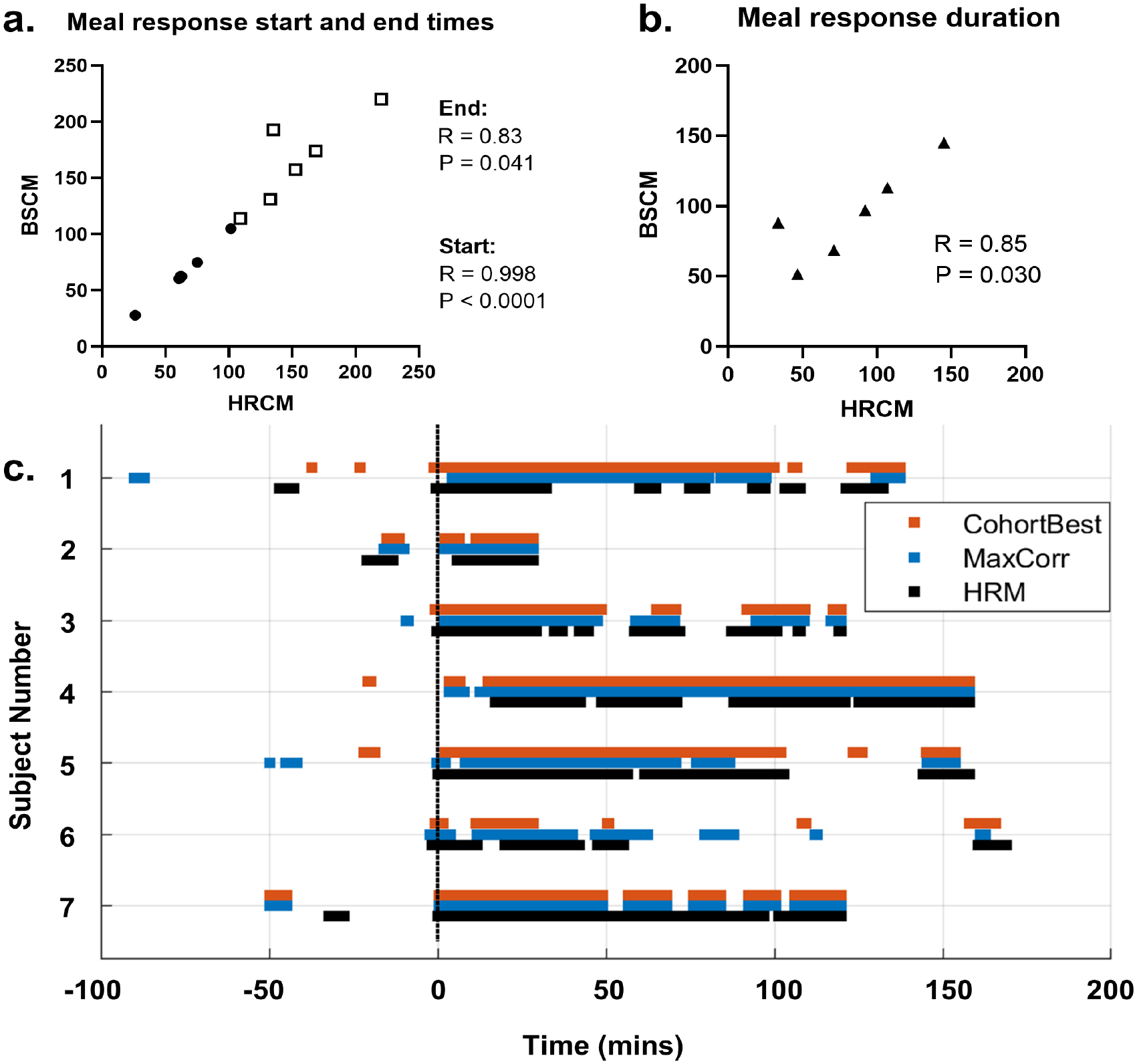
Summary of CMP active time match between HRCM and BSCM. **(a)** High level of correlation observed for both start and end times of the meal responses; **(b)** Meal response duration (meal response end time - start time) was also well correlated. Values are in units of minutes. **(c)** Suprathreshold Activity Lines show a high level of agreement in active vs quiescent periods for all subjects. Black trace = HRCM; Blue = highest correlation BSCM; Red = cohort-wide optimum parameter combination BSCM trace. Meal times are aligned at t = 0 min.

Activity lines (Fig. 4c) were auto-generated by establishing a MI level threshold (2.5 standard deviations above pre-meal MI levels) above which the colon is deemed to be active as per previous work^29^. ‘Similarity scores’ quantified the proportion of time in agreement over the course of the study (0 - no agreement; 100 = perfect agreement across the whole study period). For the ‘best’ BSCM trace (black [HRCM] vs. blue traces) the similarity score was 78.0 *±* 4.8 (median = 80.2. range = 69.1 - 82.4). For the cohort-wide best analysis combination (black vs. red [parameter set 23]) the similarity score was 78.2 *±* 8.0 (median = 80.2; range = 61.7 - 92.8). Thus, the activity line analysis, an objective measure of active times, demonstrated an overall high concordance, further corroborating the high degree of synchronicity in HRCM vs. BSCM meal response timings based on subjective visual identification via expert manual review.

### Spatial analysis

#### Spatial correlation

Spatial figures presented by the study are made using the data resulting from the signal processing combination, number 23. This was to test the sensitivity of the ‘best overall method’ to localize CMP hot zones and remove the impact that a CMR filter may have on creating accurate heat maps. Summary figures were produced with ‘main phases’ as defined from analysis of the HRCM data. These were pre-meal and meal response periods, which were ubiquitously present, as well as quiescent and secondary activity periods. Heatmap representation of CMP activity in different phases demonstrated spatio-temporal agreement between HRCM and BSCM.

For example, Figures 5 and 6 are summaries from two recordings (subjects 5 and 6) showing when CMP activity level is at baseline on HRCM (pre-meal and quiescent periods) the BSCM heatmap exhibits a concordant general paucity of activity. When the CMP activity levels are higher in the HRCM maps during the primary meal response and the secondary activity periods, the BSCM heatmaps show a similar dynamic rise in activity. A closer examination of the BSCM heatmaps’ spectral patterns revealed two main visual signatures of CMP activity; 1) bright foci on the array that represent dominant CMP active sources that lie under the array, i.e. distal colon/RSJ (c.f. secondary activity epochs of subjects 5 and 6); and 2) diffuse, low intensity changes across a large region of the body surface map caused by CMP activities projecting from a distance onto the BSCM array (c.f. proximal colon dominant primary meal response of subject 5). As the array covers the abdomen and the colon only in part, CMPs that arise from outside the array perimeter lose signal intensity inverse to the square of the source-sensor distance. The diffuse activity observed on the array can be attributed to the spatial volume conductor effect. In periods of multifocal activities, a mix of diffuse and focal activities were appreciable on the BSCM heatmaps making localization of specific hotspots challenging (e.g., during the primary meal response periods of subjects 2, 5, 6 and 7) (see Supplementary Information: Figure S1).

**Figure 5.**
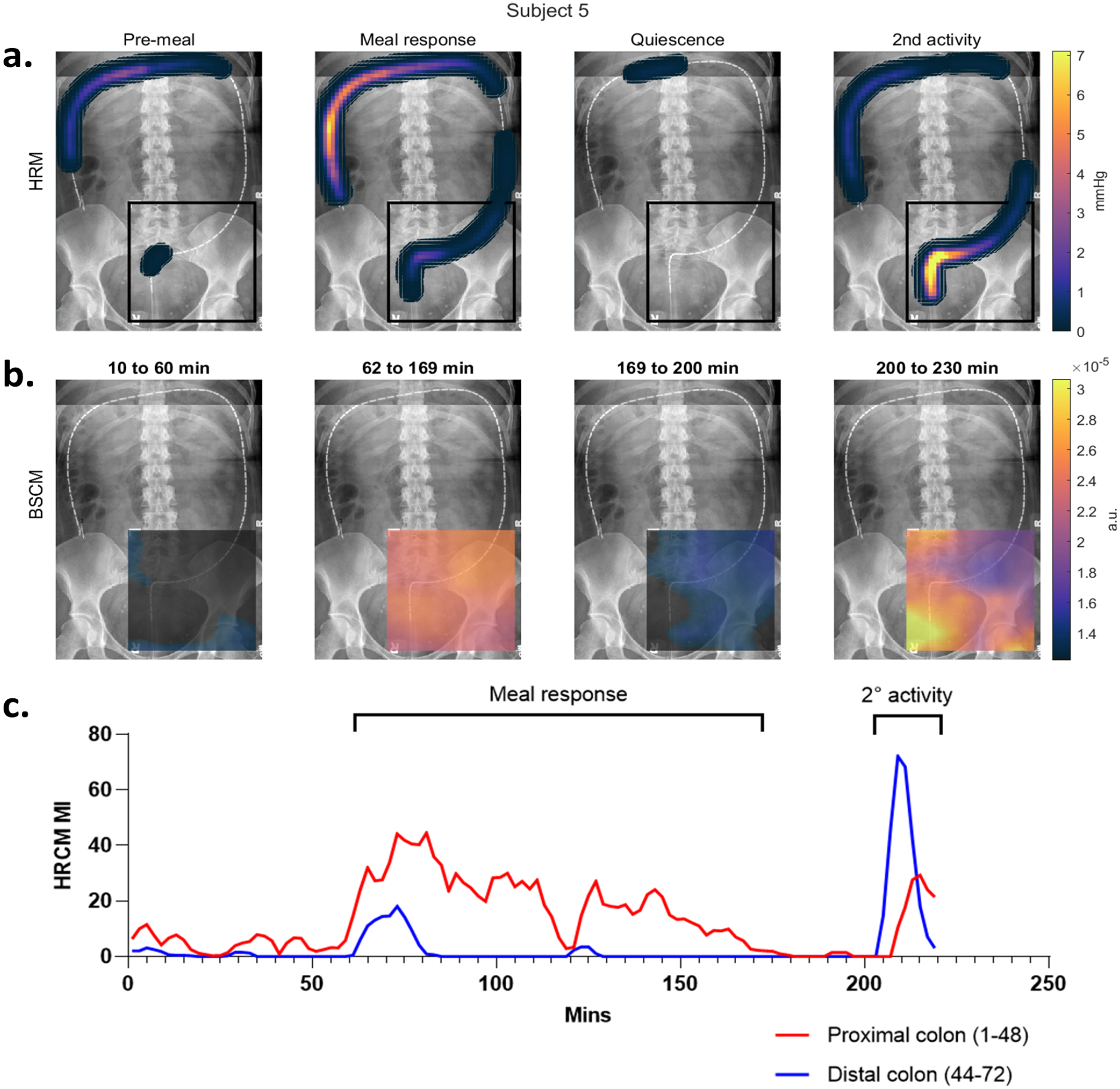
Spatial analysis of subject 5 with regional analysis of HRCM MI: Two key observations from HRCM spatial analysis **(a)** are the dominant right and transverse colon activity in the primary meal response (70-168 mins) followed by an RSJ dominant secondary period of CMP activity (200-230 min). The shift in regional dominance is mirrored by BSCM **(b)**. Distant dominant source (right/transverse colon) in the meal response period projects onto the array as a diffuse activation with weak hotspots over the RSJ and sigmoid colon. The RSJ activity in the last 20 minutes is strongly represented by a corresponding hotspot on the lower (anatomical) right corner of the array. The corresponding HRCM MI graph **(c)** shows the clear shift in regional dominance of CMP activity. The sensor numbers overlap between 44 and 48, but this was to allow for a few CMPs crossing over into the adjacent regions; no CMPs were double counted.

**Figure 6.**
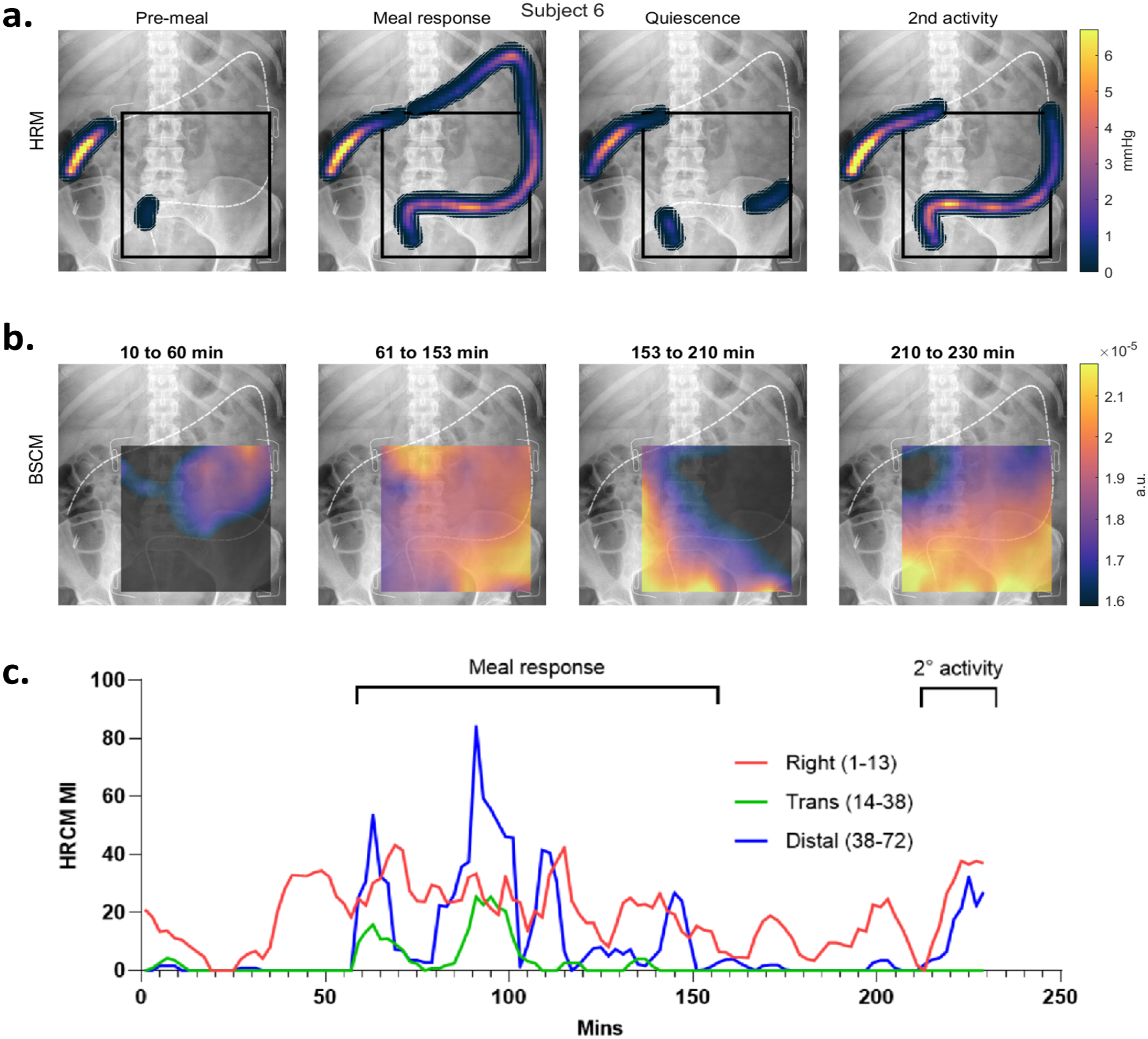
Spatial analysis of subject 6 with regional breakdown of HRCM MI: Subject 6 had a multifocal meal response involving most of the colon **(a)**. The transverse colon is closer to the BSCM array, so its activity projects a more focal impression on the top (anatomical) right side of the array during the meal response period (as seen in panel **(b)**. During the second active period (i.e. the last 20 mins), the distal transverse colon (sensors 14-38) exhibit no CMP activity (as seen in panels **(a)** and **(c)** and the spectral intensity at the top of the BSCM array is concordantly minimal.

Data was also analyzed in 10-minute epochs to examine the heatmaps in finer temporal detail (see Supplementary Information: Figure S2) however, the main spatial analysis discussions made in this study are based on the main phases.

#### Detection of dynamic shifts in the regions of dominant CMP activity on BSCM

The cases of subjects 5 and 6 also display diachronous changes to the dominant pacemaker region which altered between different phases of the recordings. This provided an opportunity to assess the sensitivity of BSCM demonstrating the changes spatially. During subject 5’s meal response period, the proximal colon is dominantly active (see Fig. 5 MI graph), followed by a 35-minute period of quiescence. In the final 30 minutes, the main activity is focused in the RSJ region. BSCM heat-maps are clearly in agreement with the manometry findings; during the meal response period, a diffuse activity pattern is observed across the array, but when the distal colon is dominant in the last 30 minutes, there is a focally bright spot localizing the CMP source to be directly over the RSJ area. In subject 6, the HRCM MI during the meal response is mainly driven by the distal colon and the right/proximal transverse colon (blue and red lines in Fig. 6 MI graph). Here, two clear active foci are observed, one hotspot on the upper left (anatomical right) array, corresponding to the proximal colon’s activity and another in the left right (anatomical left) corner that correlates to the RSJ/sigmoid regional activity. Subject 6’s transverse colon hangs low, as opposed to subject 5’s, thus the proximal colon’s activity projects a brighter distinct focus onto the array. In the last 20 minutes of the recording, the transverse colon (directly above the array) is inactive while the distal colon remains strongly active, thus the singularly active distal colon region of activity is more clearly appreciable on the BSCM heatmap.

### Participant survey

All participants preferred BSCM to HRCM. All participants who underwent both concurrently commented that BSCM was less invasive, easier, and quicker. Significantly less discomfort was incurred with BSCM (median 6.5/10 with HRCM vs. 1/10 with BSCM; *p* = 0.0005) (see Fig. 7). Only one participant mentioned that removal of the BSCM array caused discomfort as they had sensitive skin but would still prefer BSCM over HRCM. Four participants added that not requiring bowel prep was an important factor in choosing BSCM to be their preferred choice of investigation. Two participants expressed that the potential for BSCM to be undertaken out of the hospital setting without sedation and causing less disruption to work life (due to bowel prep and hospital day admission) were potential advantages.

**Figure 7.**
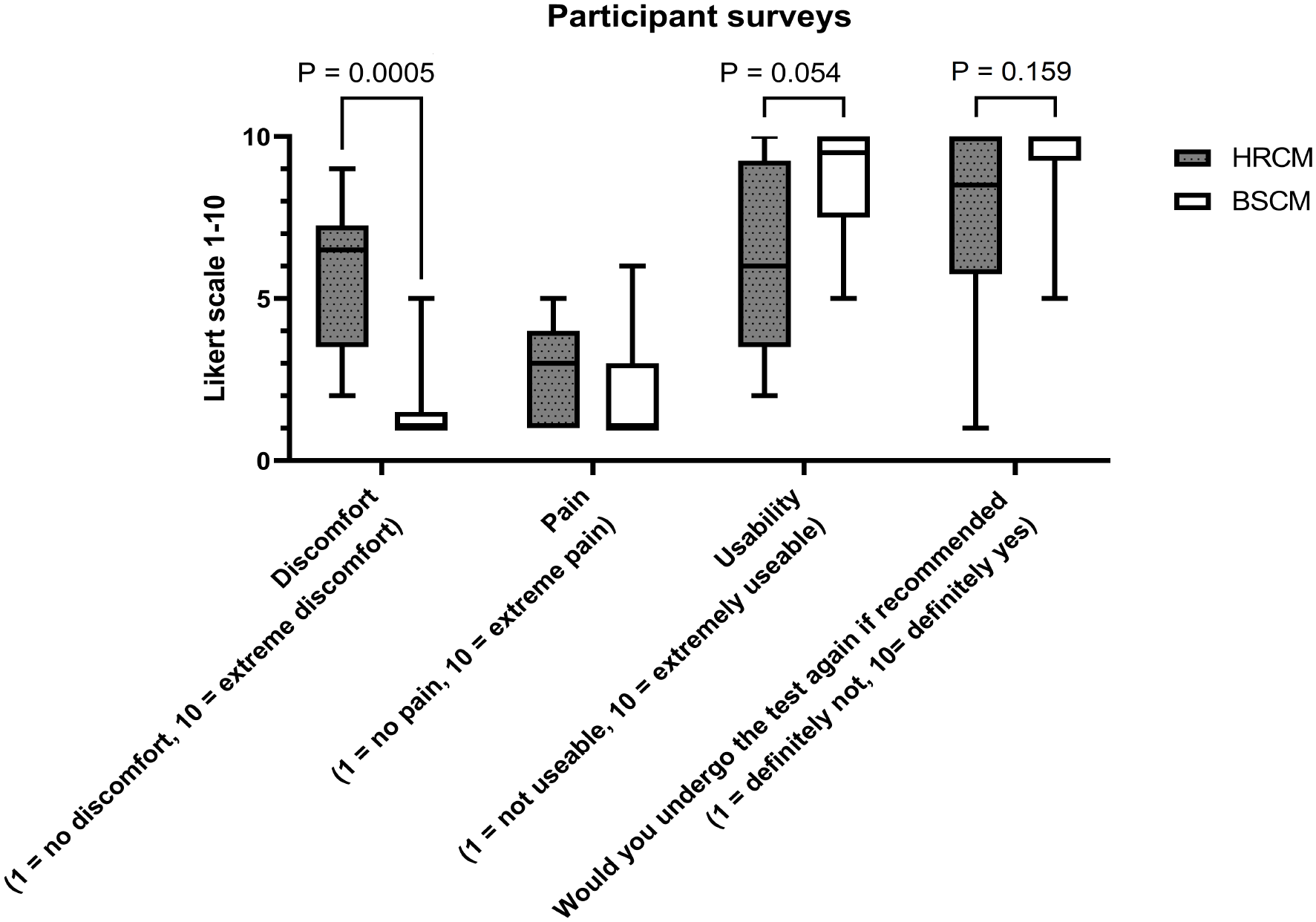
Box plot graph of the participants’ numerically reported outcomes: Non-invasive BSCM was found to be significantly more comfortable than HRCM. A trend was found favoring BSCM in outcomes regarding usability and acquiescence to repeated testing. Neither investigation inflicted significant levels of pain on the participants.

## Discussion

This study has validated BSCM as a non-invasive technique against HRCM for detecting colonic CMP activity from body surface electrical recordings. BSCM’s significantly updated CWT-based signal processing pipeline, informed from a rich set of colonic physiology analyses via HRCM, was crucial for performing temporal correlation and spatial map analyses. The study presents clear evidence that BSCM is sensitive and specific to colonic CMPs, exhibiting a high degree of spatiotemporal correlation with HRCM analyses.

As HRCM and BSCM are fundamentally different measurement modalities, motility indices were used for quantitative comparison. Motility index has been described by a number of previous manometry studies to characterize the activity density in a given epoch using an area under the curve (AUC) approach^40–45^, the definition of which has not been standardized for interpreting high resolution data. In the current study, HRCM MI was defined as a product of the 3 key HRCM outputs (number of CMPs, mean amplitude, and propagation length). The current HRCM MI metric cannot differentiate the CMP activities of lower frequency (number of CMPs per unit time) with longer distances versus higher frequency with shorter lengths in an analysis window. The BSCM MI correlation should be better in the former case but may be mismatched in the latter. An important limitation of the BSCM technique is that the directions of wave propagation cannot yet reliably be ascertained due to the complex and variable anatomy of the colon. Traditionally, HRCM studies have reported a mixture of ante- and retrograde CMP activities^34,46^, with directionality likely to play a key role in storage and organization of bowel contents, controlling passage of contents to the rectum, and the maintenance of continence^10,12,34,47^.

The bandpass filter frequency range selected for the BSCM analysis method was found to be the single most significant parameter for identifying the colonic electrophysiological signal referenced to HRCM. There was only one case (subject 5) for which correlation improved slightly with a lower frequency range approach (2-8 cpm), likely owing to the fact that a single region/single pacemaker was predominantly active at any one time, with an intrinsic frequency range of 2-4 cpm (see Fig. 3). It is worth noting that the high frequency analyses also performed well for this case. The overall superior performance of the 4-10 cpm bandwidth may be attributed to the following three factors. First, the high intrinsic frequencies in this range were observed in some cases (notably in subjects 3, 4 and 7; see Fig. 3). Further analysis of HRCM frequency distributions revealed that the largest post-vs pre-meal distribution changes occur in frequency bands > 3 cpm (see Supplementary Information: Figure S3). Second, the superposition of multiple CMP sources generates multiphasic waveforms manifesting as higher frequencies (see Supplementary Information: Table S1). For instance, independent colonic sources generating CMPs in the 2-6 cpm range, but out of phase by 10 seconds, may result in a strong *>* 6 cpm frequency (i.e., sequential frequency). Lastly, higher frequency bandwidth screens out gastric slow wave activity most efficiently. To summarize, based on the current study’s findings, the frequency range 4-10 cpm, artifact windowing, and no CMR (combination 23) was the most robust combination, and may provide a suitable analysis pipeline for future BSCM studies when HRCM referential data is absent. A caveat to the generalization of the study’s results in BSCM only studies is that the effects of the unnatural conditioning of the colon prior to recording (bowel preparation and endoscopic insertion of the HRCM catheter) on CMP activity remain to be investigated.

It is also worth noting that the optimal colonic filter bandwidth substantially differs from the 0.6-6 cpm previously validated for detecting gastric activity using BSGM^28^, which may be useful for dual colonic and gastric monitoring in future. Whereas, gastric slow waves usually propagate at a stable frequency with minimal variance (3.04 cpm; reference interval: 2.65-3.35 cpm)^48,49^48,49, the colon’s CMP frequency range is broader with a more intermittent/sporadic temporal activity profile^31^. Another major difference is that the stomach normally has a single dominant pacemaker, but the colon has multiple independent and simultaneously active regions. While others have reported a colonic frequency range measured on the body surface of 12 to 20 cpm^50,51^, we have only observed frequencies up to a maximum of about 12 cpm from HRCM analysis, which is in keeping with the large majority of manometry studies^31,33,34,52^. Also, our data clearly show that the observed frequency range from the body surface is not always directly congruent with actual colonic frequency range (due to summation effects), yet it still remains below 12 cpm. Care must be taken to differentiate colonic CMPs vs. respiratory artifacts in the 12 to 20 cpm frequency band.

The spatial mapping of the case studies presented herein illustrate for the first time that focal regions of activity can be approximately localized using the BSCM techniques. When a singular dominant region of the colon is present two observations are made: activities occurring directly beneath the array appear as focal hotspots, and activities outside of the array’s area are perceived as diffuse and broad low level of activity sensed across a large region of the array edge. Current study’s analysis also indicates that simultaneous multifocal activities lead to more diffuse heat maps that make localization challenging. This issue could be mitigated with a larger area BSCM array that would overlay the entire colon, thus CMP hotspots may be visualized with higher accuracy and focus; however, the current array setup used in this study is already a significant manufacturing challenge. More studies in the future that display dominance of meal response in different, singular regions in the colon, would be instrumental for estimating the expected contribution from different colonic sources projecting onto the spatial map.

Another limitation of the current spatial mapping technique is the assumption that electrical activities would project directly anteriorly and interpret an inherently 3-D biophysical activity using a 2-D single X-ray image. BSCM with cross-sectional imaging would likely improve accuracy in interpreting spatial patterns with a better appreciation of the variably convoluted course of the colonic tract and the abdominal wall’s shape and depth, but poses logistical, cost, and radiation challenges.

In the current study, the whole array’s data were used to derive the BSCM motility index which correlated well with manometry MI output from distal half to nearly full lengths of the colon. Many translational manometry studies have only quantified the CMP activities arising from the distal half of the colon/rectosigmoid region and it remains unclear what role proximal colon CMPs may play in function^10,12,17,34^. Localization and analysis of specific regional activities are easier with HRCM data by selecting only sensors that correlate either anatomically or physiologically with the colon region in question. By contrast However, regional analysis with BSCM is challenging due to several factors. First, the BSCM array only covers a limited segment of the colon directly, thus it is difficult to ascertain the exact source of electrical activities arising from outside the array covered area (i.e., proximal or transverse colon). In addition, due to the position of the ground and reference electrodes, on the right side (anatomical) of the array, the right colon’s CMP activity injects a common mode signal into the rest of the measurement electrodes with variable intensity depending on distance from the active current sources. In some cases (subjects 4 and 6) CMR appeared to be beneficial in correcting a predominant activity pattern observed via HRCM in the ascending colon, beneath the ground and reference electrode. However, application of CMR worsened the correlation in 2 cases where simultaneous multifocal activity was observed (subjects 5 and 7).

One limitation of this study is the small cohort size, which reflects the highly technical and challenging experimental technique which also poses difficulties to patient recruitment and throughput owing to the invasiveness of manometry, procedural stressors, and time factors, as well as COVID-related mandatory restrictions during the study period. However, together the data set is rich in that every case provided sufficient physiological and anatomical variations (frequency, regional activities, colonic anatomy, and manometer insertion depth) to inform an analysis pipeline of strong correlation which could be applied in future BSCM studies.

Although HRCM served as the ground truth in this study, it must be recognized that HRCM has a limited scope based on the extent to which the catheter is inserted (the manometer could not reach the right colon in 3 out of 7 cases). In contrast, BSCM electrodes detect a superposition of all activities, with weighted intensities dependent on source-sensor distances. For example, subject 1 had less than half the colon’s length measured by the manometer resulting in the biggest discrepancy in meal response correlations to BSCM. The data from the full colon manometry studies show that the proximal colon invariably switches on during the meal response period and BSCM heatmap analysis of subject 1 also suggests that the proximal colon remains active during the quiescent period (see Supplementary information: Figure S2).

In summary, this study has demonstrated three different metrics with validation that the resulting signal source from BSCM analysis is reliably of colonic origin: 1) motility index correlations, 2) meal response synchronicity, and 3) spatial hotspot analysis. CMP hypoactivity or hyperactivity has been associated with the development of low anterior resection syndrome (LARS)^17^, fecal incontinence^8,12^, postoperative ileus^10,53^, and irritable bowel syndrome (IBS)^13^, such that these newly validated BSCM biomarkers could serve to guide clinical therapies. This is particularly significant because while HRCM remains an important research tool, it is not widely available, relatively invasive, and its data is time-consuming and complex to analyze. BSCM, on the other hand, may be performed without anesthesia nor endoscopy, and interrogates the colon in its physiologically natural/unprepped state for longer durations. BSCM, as validated in this study, therefore has significant potential to generate a meaningful mechanistic understanding of functional disorders and guide clinical therapies.

## Data Availability

The data generated in this work will be made available upon reasonable request.

## Acknowledgments

We thank all participants in this study cohort. This project was financially supported by CSSANZ with a Foundation Grant and HRC. SHBS is a recipient of the New Zealand Research Scholarship from RACS. JCE was supported by a Washington and Lee University Lenfest Sabbatical Fellowship.We acknowledge the Colorectal Surgical Society of Australia and New Zealand (CSSANZ), Health Research Council of New Zealand (HRC) and the Royal Australasian College of Surgeons (RACS) for supporting this research. Thank you to both Phil Dinning (Flinders University, Adelaide, Australia) and Nira Paskaranandavadivel (Auckland Bioengineering Institute, University of Auckland, Auckland, New Zealand) for their early input on this study.

## Author contributions statement

SHBS: Conceptualization, methodology, formal analysis, validation, investigation, data curation, writing–original draft, visualization, and funding acquisition. CW: Methodology and data curation. TD: Methodology and data curation. DR: Investigation (endoscopy) and resources. AG: Editing. SC: Formal analysis and editing. Ian Bissett: Supervision, writing–review and editing. GOG: Supervision, writing–review and editing, and funding acquisition. JCE: Conceptualization, methodology, formal analysis, validation, formal analysis, data curation, and writing–original draft. All authors reviewed the manuscript.

## Additional information

### Data Accession

The data generated in this work will be made available upon reasonable request; please contact the corresopnding author.

