## Supplemental Material for "Validation of body surface colonic mapping against high resolution colonic manometry: a novel non-invasive tool for evaluation of colonic motility"

### TABLE OF CONTENTS

| | | |
|---|---|---|
| **Table S1:** | Frequency proportions - intrinsic vs. sequential frequency | Page 2 |
| **Figure S1:** | Spatial analysis: All subjects; major phases | Page 4 |
| **Figure S2:** | Spatial analysis: All subjects; 10 minute epochs | Page 8 |
| **Figure S3:** | Analysis of HRCM frequency distributions pre- and post-meal | Page 12 |

**Table S1: CMP activity proportions per frequency bandwidth - intrinsic vs. sequential frequency.**

| | **Intrinsic frequency** | | **Sequential frequency** | |
|---|---|---|---|---|
| | **2-4 cpm** | **4-10 cpm** | **2-4 cpm** | **4-10 cpm** |
| **Subject 1** | 48.27 | 34.52 | 20.21 | 49.20 |
| **Subject 2** | 54.65 | 23.68 | 36.18 | 42.76 |
| **Subject 3** | 47.92 | 34.30 | 34.72 | 49.46 |
| **Subject 4** | 39.00 | 43.09 | 14.23 | 55.55 |
| **Subject 5** | 51.71 | 24.03 | 31.48 | 42.43 |
| **Subject 6** | 44.85 | 34.58 | 33.71 | 42.70 |
| **Subject 7** | 38.47 | 38.52 | 14.74 | 47.14 |
| Mean | 46.41 | 33.25 | 26.47 | 47.03 |
| Std | 6.08 | 7.14 | 9.72 | 4.85 |
| Median | 47.92 | 34.52 | 31.48 | 47.14 |

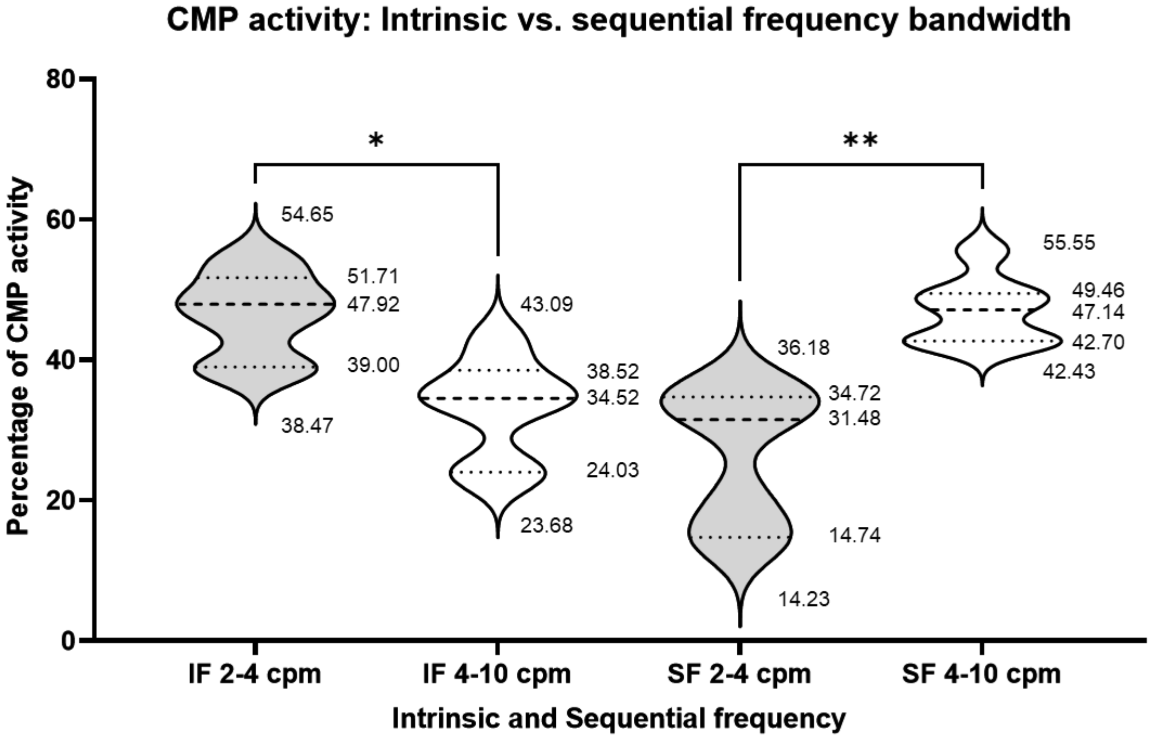

**Table S1: CMP activity proportions per frequency bandwidth - intrinsic vs. sequential frequency:** Frequency was analyzed in two ways from the manometry data. *Intrinsic frequency* is the rate of CMP activity detected on each manometer independent of other sensor data, thus representing the actual physiological frequency of the CMPs. *Sequential frequency* is the frequency of all multifocal CMP activities combined; i.e. all active colonic segments are viewed as one single active organ (when only one region is active, the frequency will equal intrinsic frequency). Intrinsic frequency of the colon is predominantly around 2-4 cpm, however, the proportions of CMP activity seen via the sequential frequency bandwidths are the opposite and the 4-10 cpm range becomes the dominant bandwidth. The sum of the two bandwidth values do not add up to 100 as the proportions of minor frequency bandwidths below 2 cpm and above 10 cpm are not shown.

**Figure S2. Spatial analysis: All subjects; major phases**

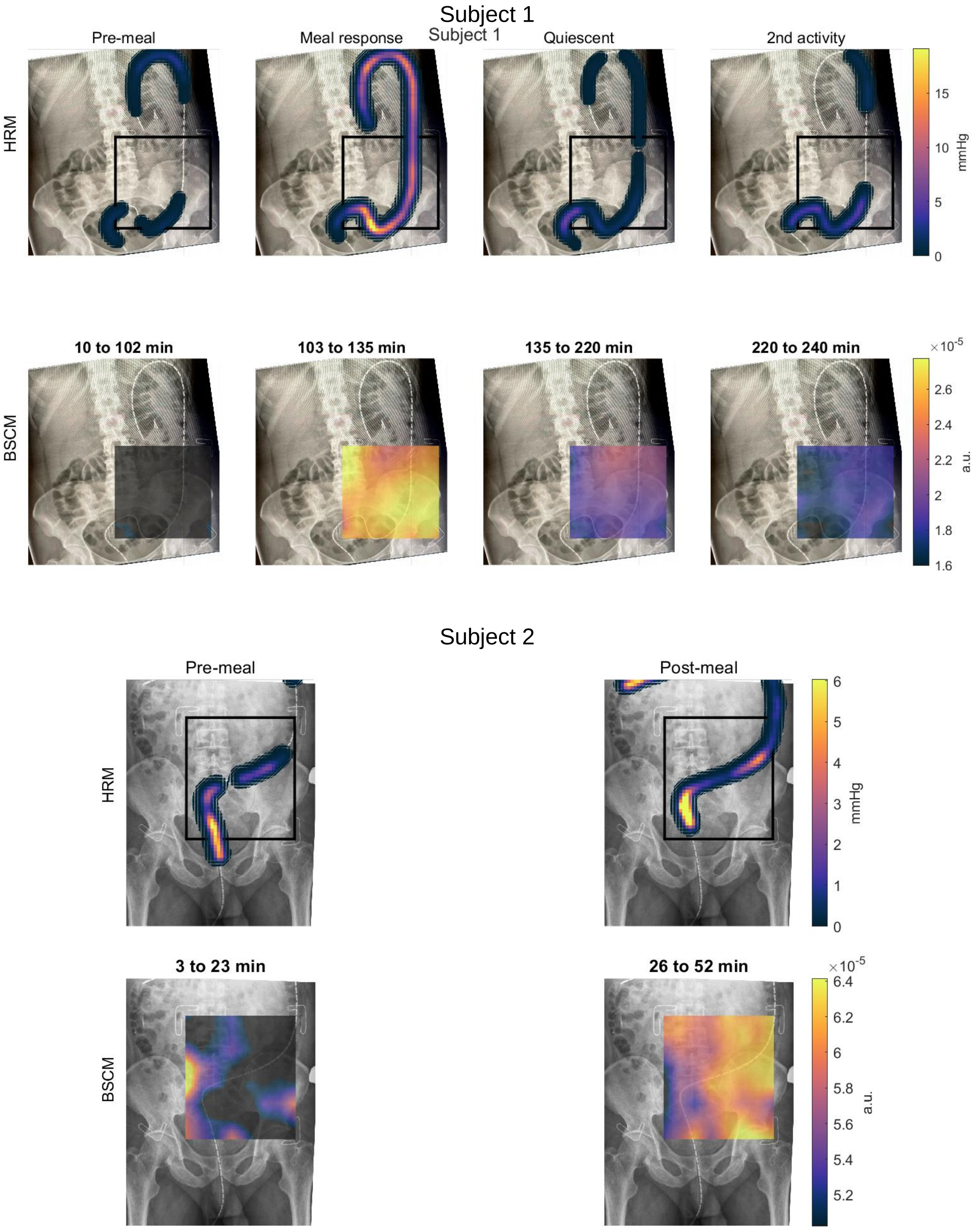

Subject 3

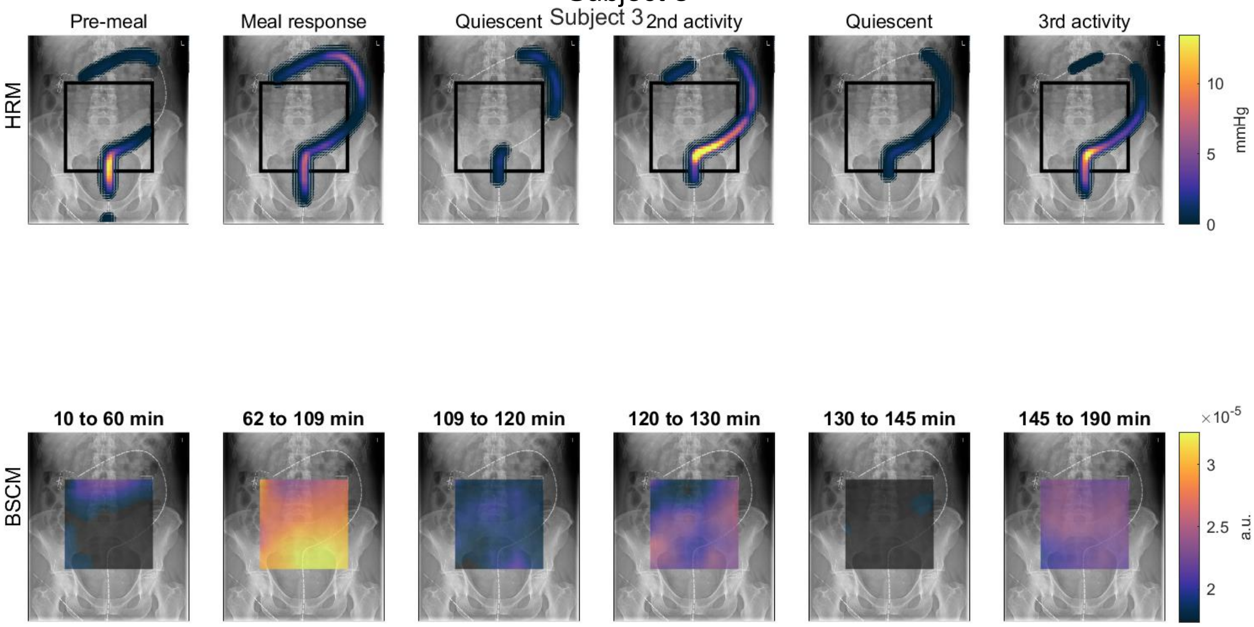

Subject 4

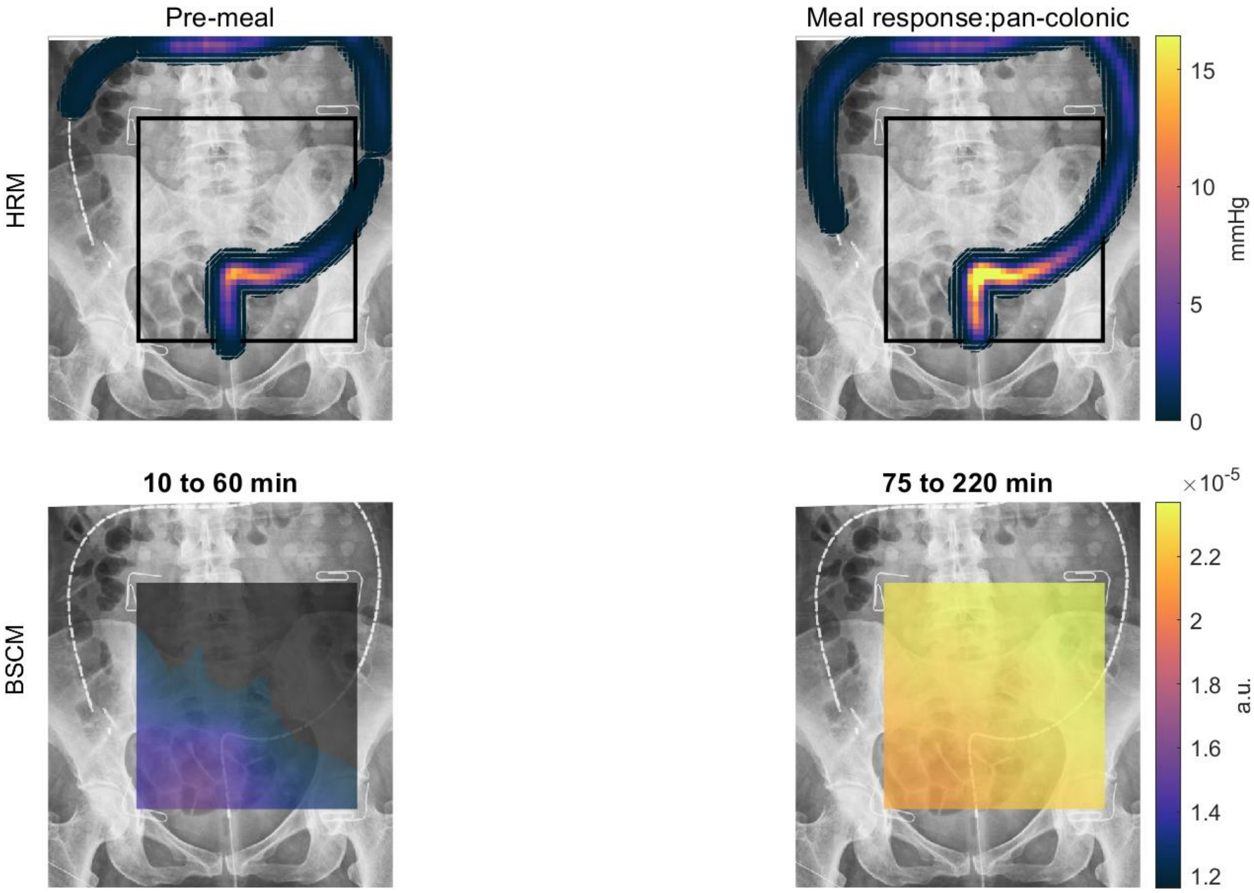

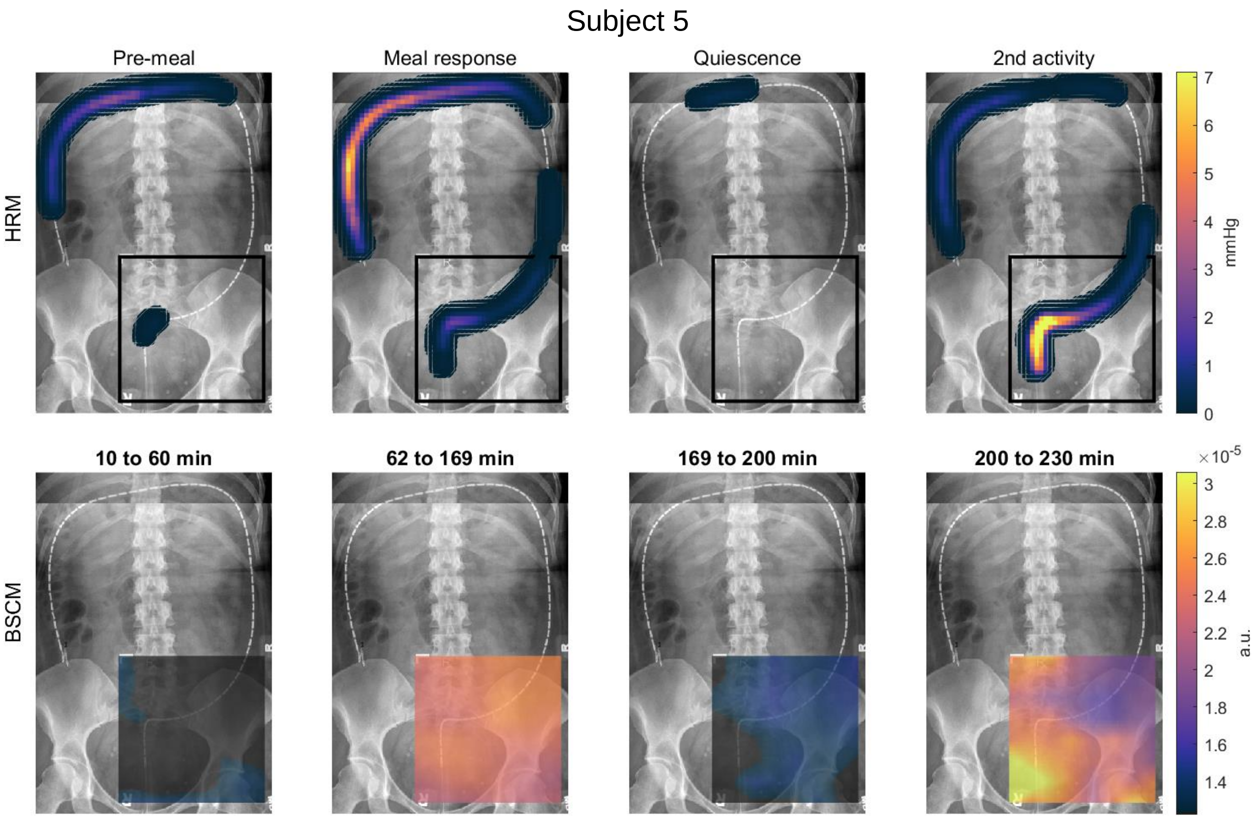
Subject 5
Pre-meal
Meal response
Quiescence
2nd activity
HRM
mmHg
10 to 60 min
62 to 169 min
169 to 200 min
200 to 230 min
BSCM
a.u.

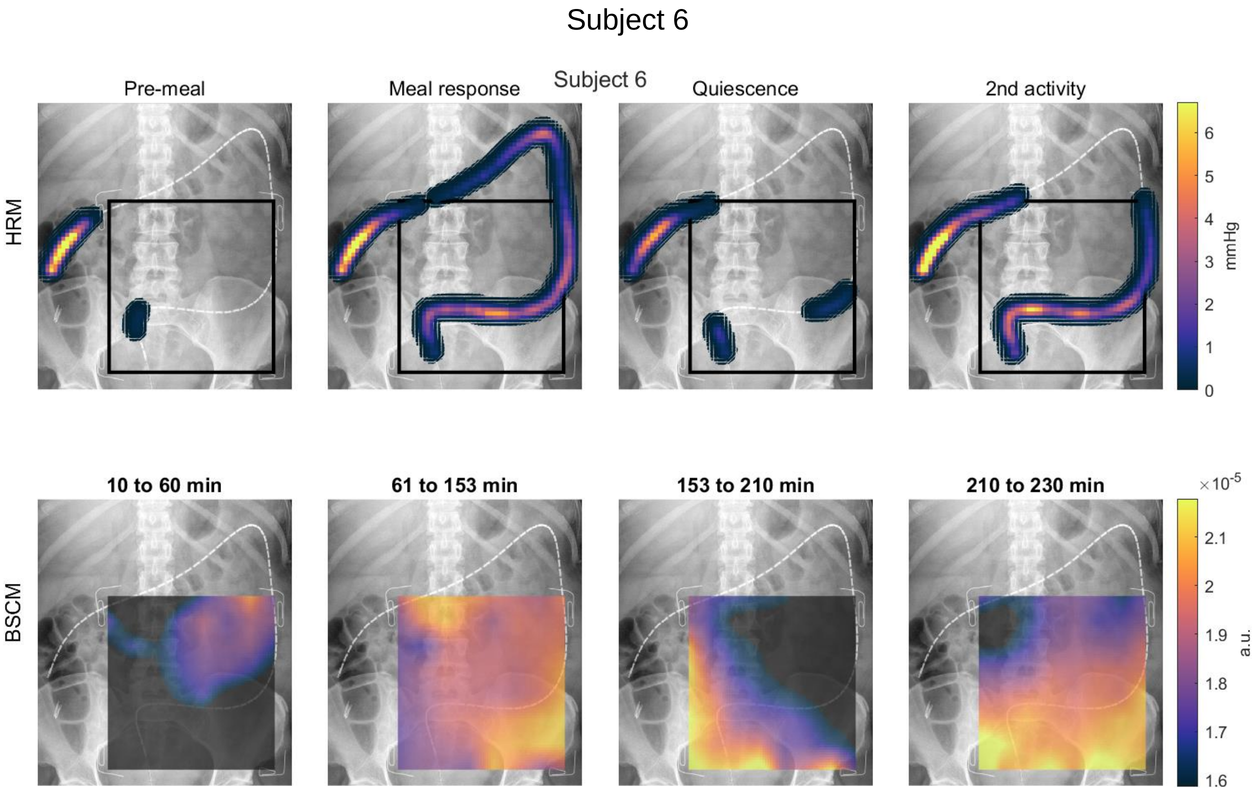
Subject 6
Subject 6
Pre-meal
Meal response
Quiescence
2nd activity
HRM
mmHg
10 to 60 min
61 to 153 min
153 to 210 min
210 to 230 min
BSCM
a.u.

Subject 7

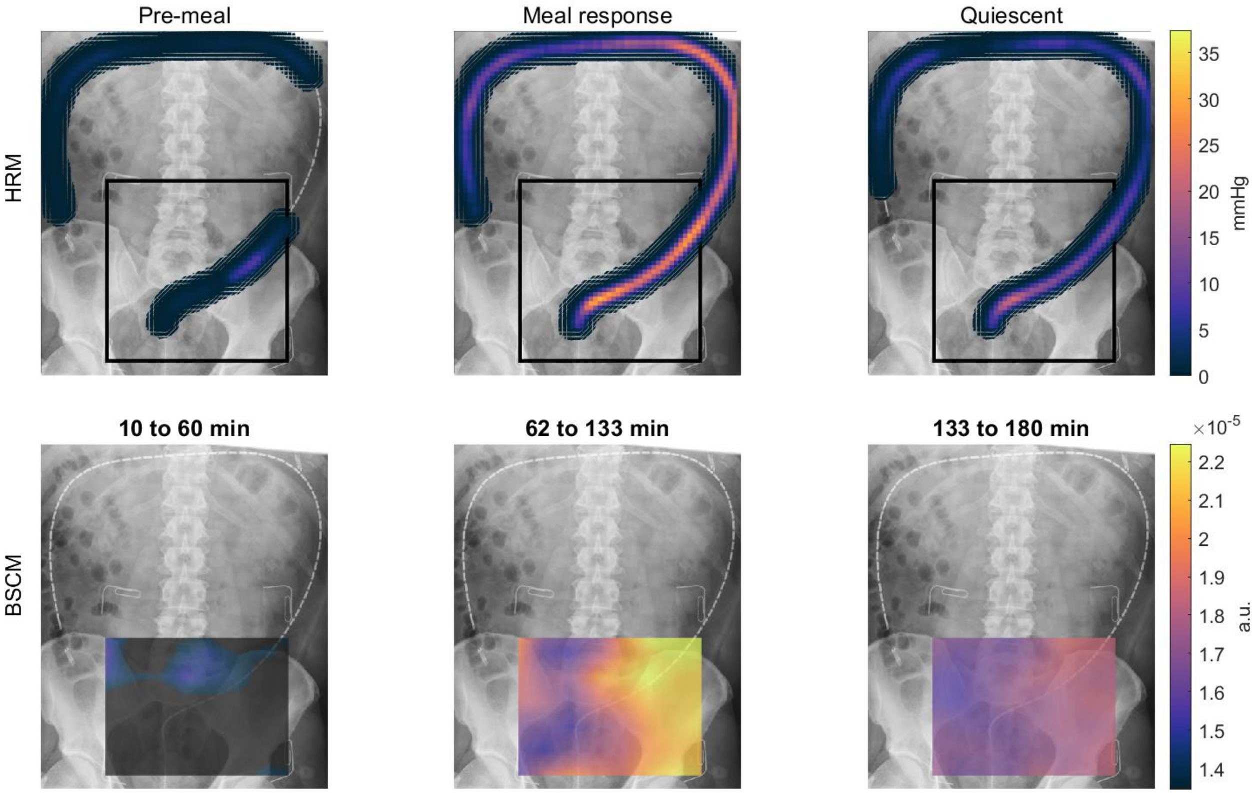

Pre-meal
Meal response
Quiescent
HRM
BSCM
10 to 60 min
62 to 133 min
133 to 180 min
mmHg
0
5
10
15
20
25
30
35
a.u.
×10^-5
1.4
1.5
1.6
1.7
1.8
1.9
2
2.1
2.2

**Figure S1. Spatial analysis: All subjects; 10 minute epochs.**

Subject 1

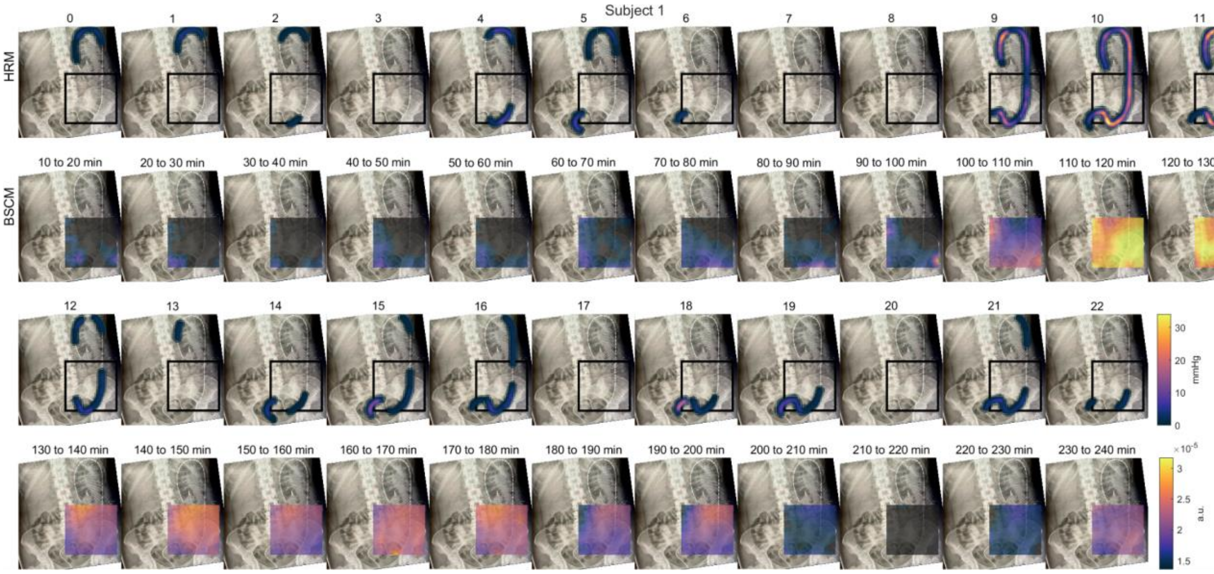

Subject 2

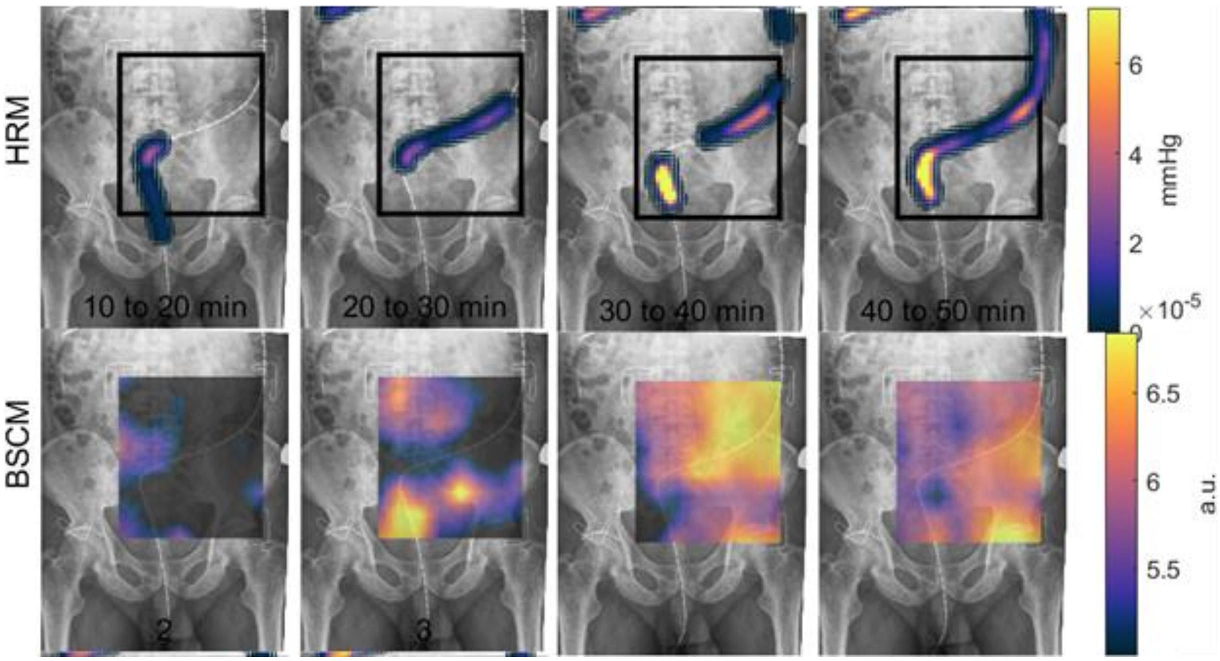

Subject 3

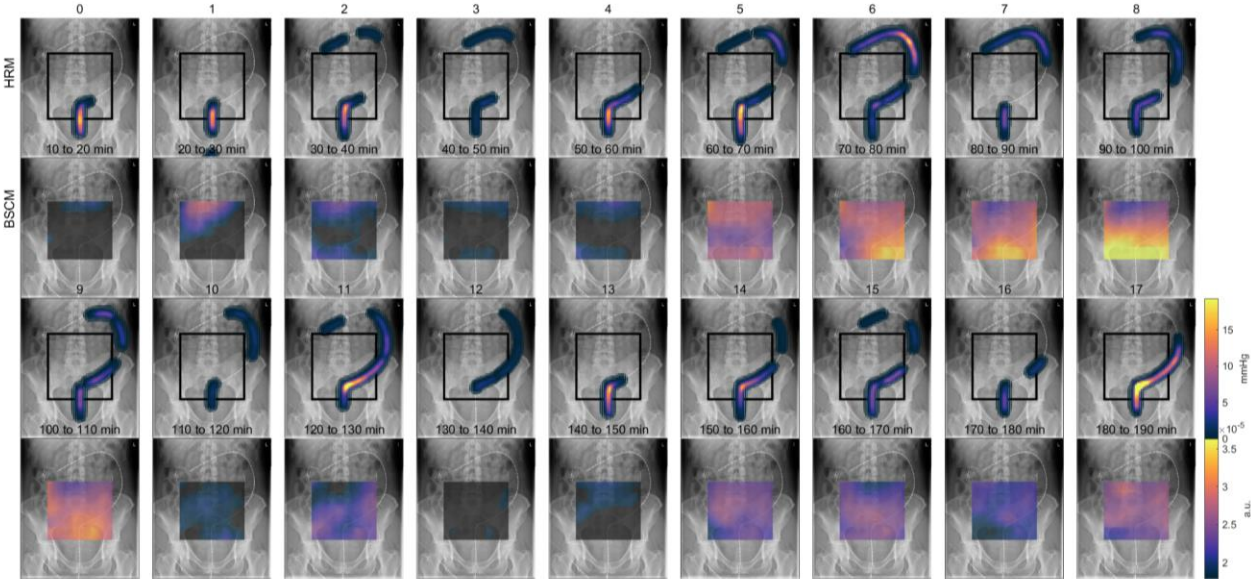

Subject 4

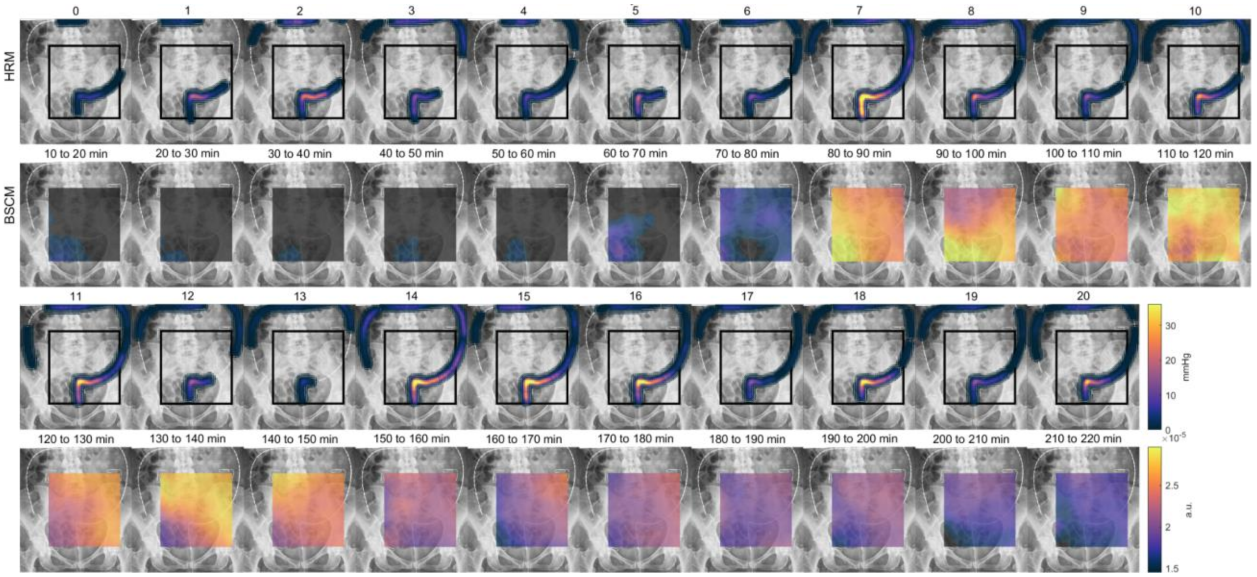

### Subject 5

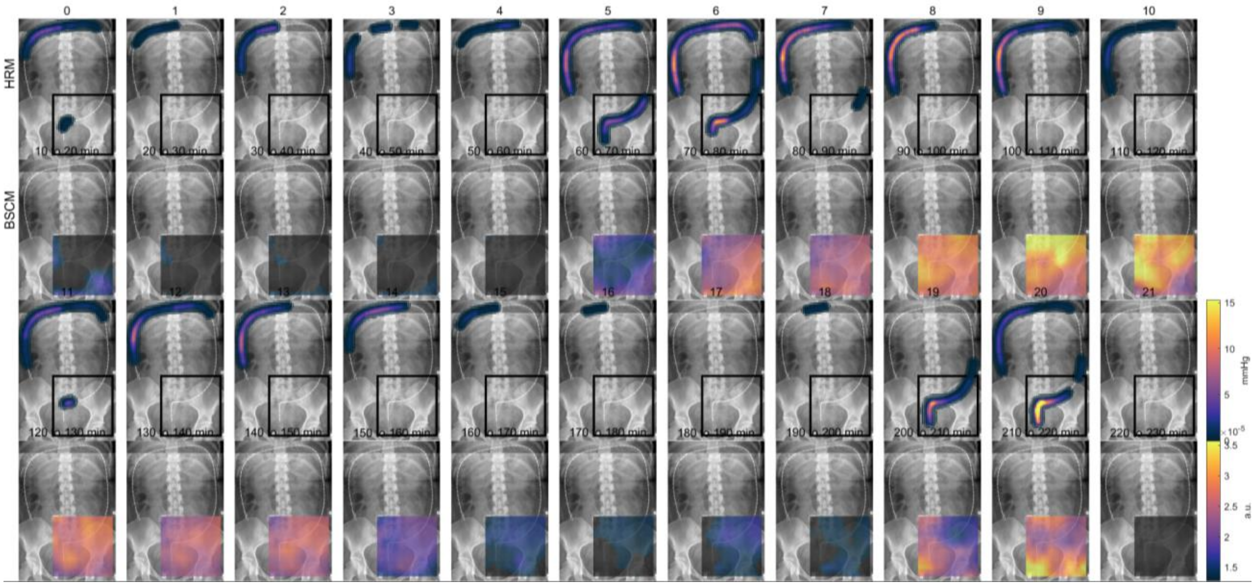

### Subject 6

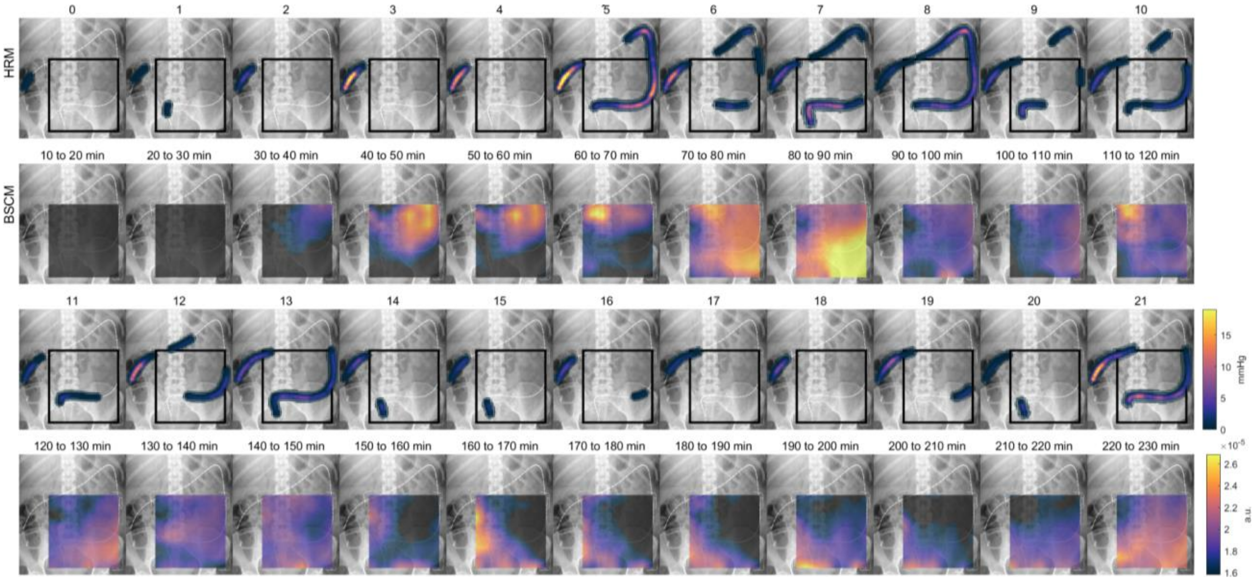

Subject 7

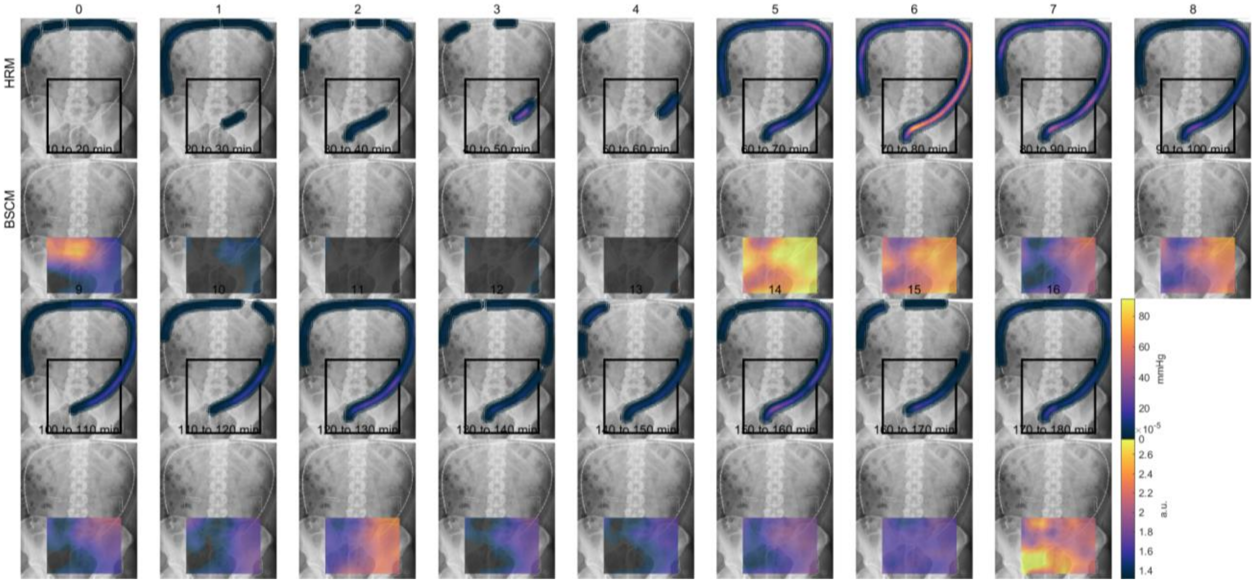

HRM
BSCM
10 to 20 min
20 to 30 min
30 to 40 min
40 to 50 min
50 to 60 min
60 to 70 min
70 to 80 min
80 to 90 min
90 to 100 min
100 to 110 min
110 to 120 min
120 to 130 min
130 to 140 min
140 to 150 min
150 to 160 min
160 to 170 min
170 to 180 min
mmHg
80
60
40
20
0
×10^-5
a.u.
2.6
2.4
2.2
2
1.8
1.6
1.4

**Figure S3. Analysis of HRCM frequency distributions pre and post-meal.**

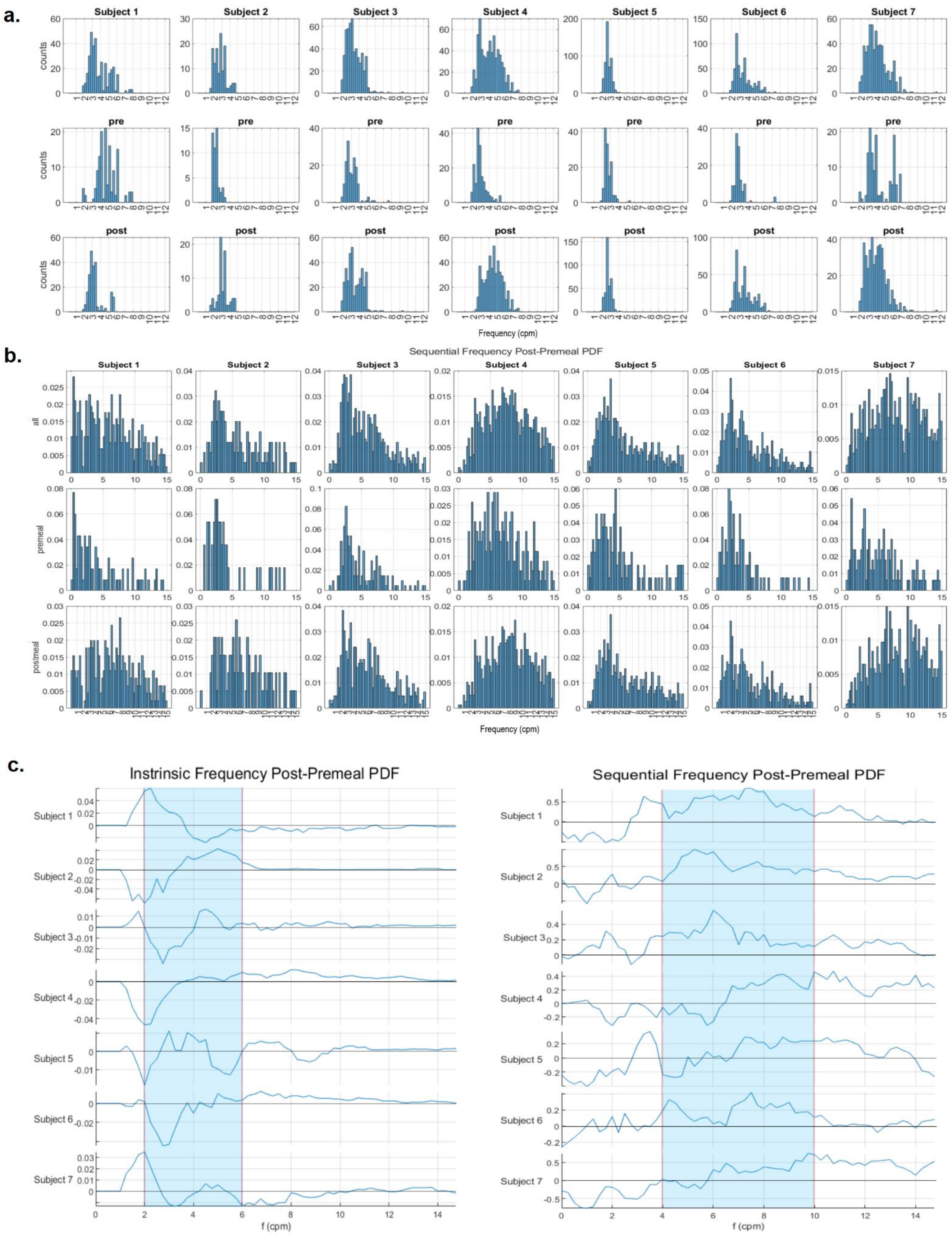

**Figure S3: Frequency distributions and changes pre and post-meal.** Histograms panels of intrinsic (**a**) and sequential (**b**) frequency distributions are split into overall (first row) distribution followed by a subanalysis of pre- (second row) and post-meal (third row) frequency distributions. Frequency is in cycles per minute (X = frequency, Y = epoch count). Graphs from panel (**c**) show the differential between pre- and post-meal intrinsic (left) and sequential (right) frequencies for each subject. Biggest rises post-meal are noted in the 2-6 cpm range for the intrinsic frequency and in the 4-10 cpm range for the sequential frequency.
